# Quantifying Systematic Bias in Observational Causal Estimates Using Negative Controls: The Bias Attribution Fraction

**DOI:** 10.64898/2026.10.08.26365019

**Authors:** Zeng Zhechun, Wang Jinwen, Zuo Huijuan, Shu Lixia

## Abstract

**Importance:** Observational studies dominate evidence in cardiovascular outcomes research, yet no widely adopted reporting standard requires authors to disclose *how much* of an observed association is attributable to systematic error. The negative-control calibration framework of the Observational Health Data Sciences and Informatics consortium returns a binary answer: a calibrated P-value either crosses 0.05 or it does not. That answer conflates “is there a signal” with “is the signal credible.”

**Objective:** To define the Bias Attribution Fraction (BAF), a continuous estimator in [0, 1] that quantifies the fraction of an observed log relative risk that is attributable to systematic bias, together with its 95% credible interval, and to evaluate whether reporting calibrated P-value plus BAF plus interval width produces more defensible decisions than reporting the calibrated P-value alone.

**Design:** Three-part study: (i) metric definition and interval construction; (ii) Monte Carlo simulation across 1920 conditions × 1000 repetitions = 1,920,000 simulated target estimates (a primary grid plus a sign-flipped companion grid of identical size, pooled for all analyses), with truth known by construction; (iii) two-arm beta-blocker target trial emulation in MIMIC IV, adhering to the TARGET guideline, with targeted maximum likelihood estimation (TMLE) and 12 negative-control outcomes.

**Setting:** MIMIC IV (critical care) and MIMIC IV ECG, 2008 to 2019.

**Participants:** Adults with a first intensive care unit stay and a recorded 12-lead ECG.

**Exposures:** Two clinical questions are evaluated: (1) reaching at least 50% of guideline-recommended target dose of an index beta-blocker within 7 days of admission, sustained to 12 months, versus lower or no exposure; (2) receiving guideline-directed medical therapy (GDMT) with at least two of three foundational drug classes at adequate dose for 12 months versus fewer.

**Main Outcomes and Measures:** One-year all-cause mortality. Primary analytic outputs: (i) the calibrated P-value from a negative-control empirical null distribution with 12 outcomes; (ii) BAF̂ with a Markov chain Monte Carlo 95% credible interval; (iii) a verdict drawn from a truth-calibrated lookup table mapping the BAF̂ × interval-width cell to a probability of being bias-dominated.

**Results:** Across 1,920,000 simulated repetitions, the calibrated P-value alone yielded a misuse rate (declared usable while truth was bias-dominated) of 36.02%. Adding the BAF point estimate reduced misuse to 16.48%, and requiring the whole credible interval of BAF to lie below 0.5, clear of the bias-dominated zone, reduced misuse to 10.16%. Requiring in addition a calibrated probability of bias dominance below 15% reduced misuse to 3.02% at the cost of declaring 22.1% of estimates usable (versus 61.8% for the calibrated P-value alone). Read from the bias-dominated view, meaning that withholding a repetition is a positive call and being truly bias-dominated is a positive truth, the BAF estimator achieved an area under the receiver operating characteristic curve of 0.922, compared with 0.787 for the calibrated P-value. In MIMIC IV, question 1 produced a calibrated P-value of 0.894 and BAF̂ = 0.875 (95% credible interval 0.385 to 0.992); the apparent one-year mortality reduction of 16.9% was entirely attributable to systematic bias and the association was not usable as effect evidence. Question 2 also failed layer 1, with a calibrated P-value of 0.1026, and BAF̂ = 0.460 placed it in the “competitive, no verdict” zone.

**Conclusions and Relevance:** Pairing the calibrated P-value with BAF and its 95% credible interval turns a binary credibility judgement into a three-channel report (signal existence, effect size, and bias fraction). Despite TARGET-adherent design, doubly robust TMLE, and 12 negative controls, both clinical questions in this study still failed to qualify as effect evidence, with question 1 demonstrating complete bias domination and BAF quantifying the systematic error that remained. The reporting standard proposed here can be applied to any observational study that includes negative controls, without requiring additional data.

## Introduction

Observational studies are the dominant source of evidence for cardiovascular effectiveness and safety questions that cannot be answered by randomized trials. They are also vulnerable to systematic error that survives the most rigorous design choices available to observational research: explicit eligibility criteria, time-zero alignment, intention-to-treat and per-protocol analyses, and modern doubly robust estimators such as targeted maximum likelihood estimation (TMLE)^1,2^. A common reaction to residual error is to require authors to perform a sensitivity analysis^3^. Sensitivity analyses ask the analyst to stipulate how much unmeasured confounding would have to exist to overturn the result, then return a qualitative judgement. They do not quantify how much of the observed association is plausibly attributable to bias.

The Observational Health Data Sciences and Informatics (OHDSI) consortium has proposed a complementary practice: estimate an empirical null distribution from a panel of negative-control outcomes, jointly estimate its center and scale, and use that distribution to produce a calibrated P-value and a calibrated effect estimate^4^. The calibrated P-value separates the question “does a signal exist after removing systematic error” from the question “how large is the effect.” That is a useful step, but it returns a binary answer for the first question and a point estimate for the second, leaving the analyst to decide, without guidance, whether the residual estimate is dominated by bias or by effect.

This manuscript develops a third quantity, the Bias Attribution Fraction (BAF), defined on the log relative risk scale as the fraction of the absolute observed association attributable to the absolute estimated systematic bias:

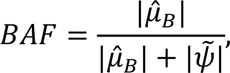

where *μ̂_B_* is the estimated center of the empirical null and *ψ̃* is the calibrated log relative risk. BAF lies in [0, 1], equals 0 when the calibrated effect absorbs the entire magnitude, and equals 1 when systematic bias absorbs the entire magnitude. The absolute values mean that one BAF is consistent with bias that inflates the effect, shrinks it toward the null, or reverses its sign, and the primary data-generating grid of Part II sets the true effect and the true systematic bias to the same sign, so the estimator and the rules are characterized in detail for the regime in which bias inflates the apparent effect. A companion grid of identical size flips the sign of the true bias, reproducing the opposing roles in which the bias shrinks the apparent effect, cancels it, or reverses it; that grid is analysed in the supplementary material (Table S15 and Table S16), its consequences for the interval and for the rules are noted in Limitations, and the two grids are pooled for every performance analysis reported below. We use BAF as the primary metric and BAF̂ and its 95% credible interval as the principal object of inference. BAF is related to the bias-to-effect ratio by *BAF* = *BER*/(1 + *BER*); BER is reported as an auxiliary summary to maintain consistency with prior reports from this program of work.

The manuscript is organised in three sequential parts. Part I defines BAF and the credible-interval construction. Part II evaluates the metric in a 1,920,000-repetition Monte Carlo simulation. Step 1 of Part II asks whether BAF̂ is an accurate estimator of BAF_*true*_ in a Bland-Altman sense. Step 2 of Part II asks whether a two-layer decision rule, calibrated P-value plus BAF̂ plus credible interval, reduces decision error relative to the calibrated P-value alone. Part III applies the calibrated rule to two clinical questions about beta-blocker therapy drawn from a target trial emulation in MIMIC IV^5–7^. The two questions are selected as a contrasting pair: question 1 (achieving ≥50% of guideline-recommended target dose) is a hypothesis that has repeatedly been reported as protective in observational studies and is a strong candidate for healthy-user bias; question 2 (GDMT with at least two of three foundational drug classes at adequate dose) is closer to a positive control with randomized-trial support. The two-arm contrast allows us to test whether the rule is informative on a case where a true effect is expected to exist.

Across all three parts the same estimator pipeline is used, and the decision thresholds used in Part III are calibrated from the Part II simulation, not from the empirical data. This separation of training and application is the central methodological discipline of the study. The case study is deliberately a maximal test: the emulation is built to the TARGET guideline, estimated with doubly robust TMLE and calibrated with 12 negative controls, asking whether systematic error survives even the strictest observational design.

Several existing approaches also probe unmeasured confounding, and BAF is designed to complement rather than replace them. Quantitative bias analysis asks the analyst to specify a bias model from external evidence, propagate it, and report a bias-adjusted estimate and interval^8,9^; it is most informative when such external evidence exists, and least informative precisely when the bias is unknown, which is the situation BAF addresses. External-adjustment and bounding approaches follow the same logic, requiring an assumed magnitude for the unmeasured confounder and returning an adjusted estimate rather than a share of the observed association^10,11^. Negative-control methods, including the empirical calibration we build on, ask whether an estimate is distinguishable from a distribution of bias estimated from the data themselves, but stop at a calibrated P-value^12,13^. BAF uses that same empirical null and reports the share of the calibrated signal attributable to bias on a scale bounded between 0 and 1.

## Methods

### Part I. The metric

Let *Y_k_*, *k* = 1, …, *K*, denote the estimated log relative risk for the *k*-th negative-control outcome in a study that estimates an analogous quantity for the target outcome *Y*. We assume

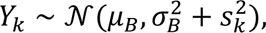

where *μ_B_* is the systematic bias center,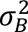 is the between-negative-control variance of the systematic bias, and 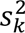 is the sampling variance of the *k*-th negative-control estimate. The pair (*μ_B_*, *σ_B_*) is jointly estimated by maximum likelihood, with *σ_B_* capped at the empirical standard deviation of the *K* estimates to preserve positive-definiteness. The calibrated P-value is then the upper tail probability of *Y* under the fitted empirical null, computed as 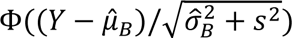 where *s*^2^ is the sampling variance of the target estimate. The calibrated log relative risk is *ψ̃* = *Y μ̂_B_*.

BAF is defined as above. Because BAF is a ratio function of two jointly estimated quantities, the delta method produces unstable inferences when *ψ̃* is near zero. We therefore propagate uncertainty rather than approximate it: a set of draws of *μ_B_* is generated from the fitted systematic error model, and each draw is passed through the same function *BAF* = |*μ_B_*|/(|*μ_B_*| + |*μ_B_*|), with the observed target estimate *Y* held at its observed value. The 2.5th and 97.5th empirical quantiles of the resulting BAF distribution form the interval, and the point estimate is the median of that same distribution, The interval is therefore a credible interval, not a confidence interval: no sampling distribution is inverted, so it carries no frequentist coverage guarantee, and its realised coverage in the simulation is reported in the Limitations. In the simulation the draws are posterior draws from the systematic error model 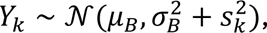 fitted by a Gibbs sampler run for 400 iterations with the first 100 discarded as warm-up, leaving M = 300 retained posterior draws, with priors 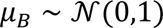 and 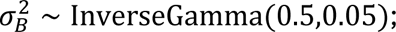 in the case study they are nonparametric bootstrap resamples of the negative-control panel, with the empirical null refitted within each resample. Both constructions yield a distribution for BAF itself, and a bootstrap percentile distribution can be read as an approximation to a posterior under an uninformative prior, so we use the single term credible interval for both. The empirical null is estimated with the EmpiricalCalibration package^4^; the BAF extension is provided by the accompanying R code archived with this manuscript.

### Part II. Simulation study

The simulation was designed to stress-test the rule across the regions of the parameter space most relevant to observational cardiovascular research. Five factors were crossed in a full factorial design within each of two half-grids that differ only in the sign of the bias center, yielding 1920 conditions in the pooled design. The factors and levels are listed in Table 1. Each condition was replicated 1000 times, for a total of 1,920,000 repetitions. The data-generating process used a logistic regression with 12 to 50 negative-control outcomes, -0.40 ≤ ψ ≤ -0.01 on the log relative risk scale, *μ_B_* from -0.40 to -0.05 and from 0.05 to 0.40 in the companion grid, and bias exchangeability held or violated at 30%. Each repetition yielded a triple consisting of a truth BAF_*true*_ ∈ [0, 1], an estimated BAF̂, and a calibrated P-value. The simulation covered 52.5% bias-dominated, 28.7% mixed, and 18.8% effect-dominated repetitions. Factor ranges were chosen to span the magnitudes typical of cardiovascular observational research: true protective effects from marginal to large, and systematic-bias centers matching the healthy-adherer effects repeatedly reported for adherence-related exposures^14,15^. Percentages describing truth composition refer to all 1,920,000 repetitions of the pooled design; every decision-rule percentage states its denominator explicitly (all repetitions, layer-1 passable repetitions, or repetitions declared usable). With this many repetitions, the Monte Carlo standard error of any reported proportion is below 0.07 percentage points. The design follows recommended practice for simulation studies^16^. All simulations and analyses were implemented in R (version 4.4.1) using the bafratio (version 0.4.0), EmpiricalCalibration (version 3.1.4), and ltmle (version 1.3.0) packages, and the Monte Carlo grids were generated with fixed random seeds (43 for the primary grid and 100,043 for the companion grid, whose random number stream is disjoint from the primary one) for reproducibility.

**Table 1.**
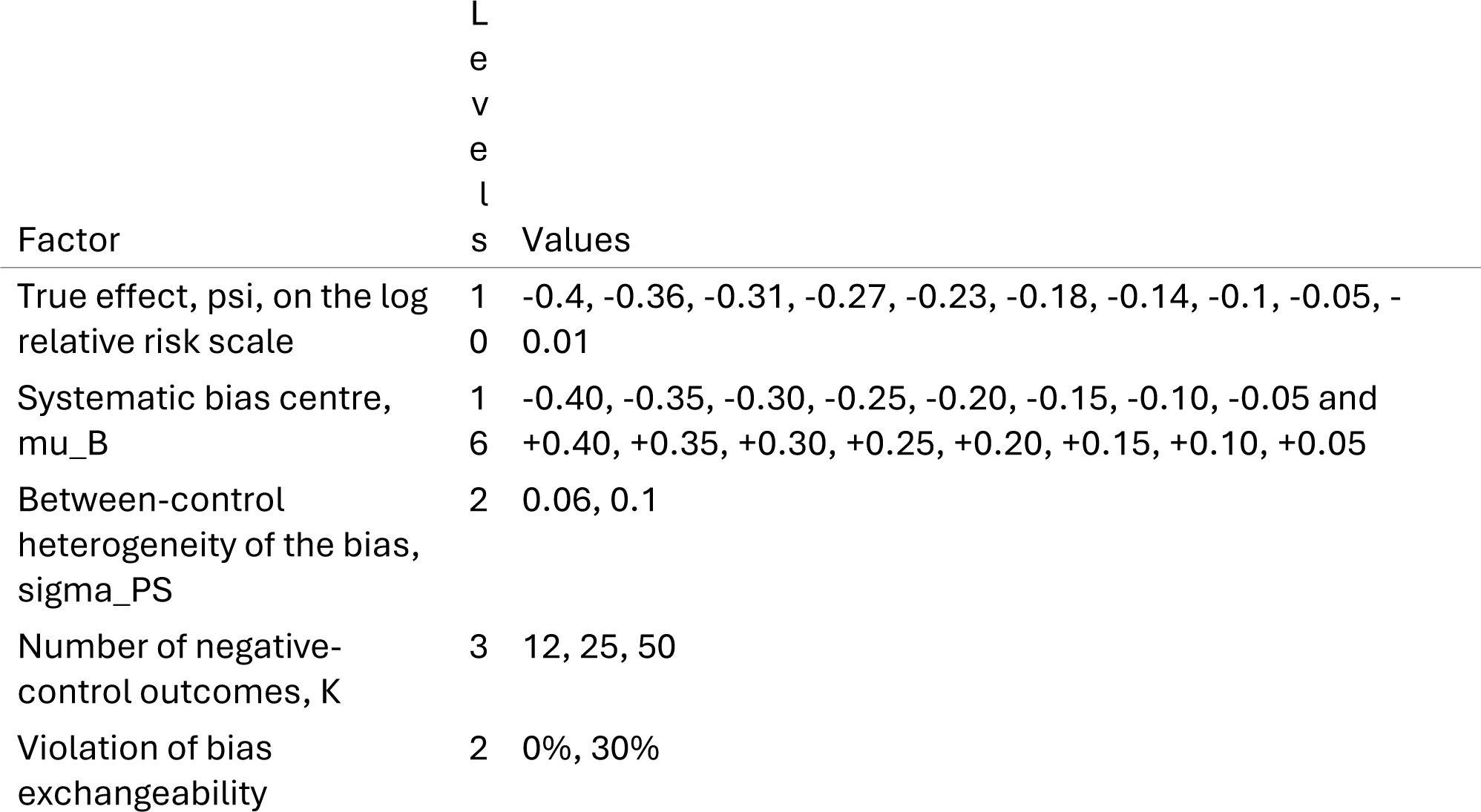
Five-factor simulation design. The five factors were crossed in a full factorial design within each of two half-grids that differ only in the sign of the bias center, giving 1920 conditions in the pooled design, and each condition was replicated 1000 times, for 1,920,000 simulated target estimates in total. The resulting composition of truth is 18.8% effect-dominated, 28.7% mixed, and 52.5% bias-dominated.

### Step 1. Bland-Altman agreement

The accuracy of BAF̂ as an estimator of BAF_*true*_ was evaluated in the classic true-value formulation: the per-repetition signed difference 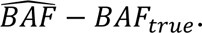 is plotted against the known truth *BAF_true_*^17^. We report the mean signed difference, the standard deviation of the difference, the 95% limits of agreement, Lin’s concordance correlation coefficient (CCC)^18^, and the proportional-bias slope (the regression of the difference on *BAF_true_*), which detects directional underestimation that grows with *BAF_true_*. The interior analysis excludes the *ψ* = -0.01 level to avoid the near-boundary pathologies that arise when the calibrated effect approaches zero; a full analysis including the boundary level is also reported. A clinical-view variant, plotting the difference against the condition-mean predicted BAF̂ that a clinician actually observes, is provided in the companion interactive explainer.

### Step 2. Two-layer decision rule

The decision rule has two layers. Layer 1 is the calibrated P-value, declaring the repetition “passable” if *p*_cal_ < 0.05. Layer 2 then operates on the subset that passed layer 1, indexing BAF̂ into a 12-bucket equal-frequency lookup table and using the bucket center together with the credible-interval half-width, classified as “narrow” or “wide” relative to the median 0.088, to read a probability of being bias-dominated. The conservative value max(*p*_BD, narrow_, *p*_BD,wise_) is used for the verdict. The four verdict zones are defined by prespecified probability thresholds of 0.15, 0.45, 0.65, corresponding to “usable as effect evidence,” “mixed, hypothesis-generating,” “competitive, no verdict,” and “not usable as effect evidence.” A second, independent rule works directly on the BAF scale, which is divided at 1/3 and 0.5 into an effect-dominated zone below 1/3, a mixed zone, and a bias-dominated zone above 0.5: it requires the whole credible interval of BAF̂ to lie below 0.5, clear of the bias-dominated zone, rather than only its point estimate, and is also evaluated as a stand-alone alternative. Nested comparisons R0 through R4 add one component at a time: R0 is uncalibrated P-value, R1 is calibrated P-value alone, R2 adds BAF̂ < 0.5, R3 adds the credible-interval requirement just described, and R4 adds the requirement that the calibrated probability of bias dominance fall below 15%.

### Part III. Beta-blocker target trial emulation

#### Data source

MIMIC IV (critical care) and MIMIC IV ECG^19,20^, covering intensive care unit stays at Beth Israel Deaconess Medical Center between 2008 and 2019. The two databases were linked at the stay level. Adults aged 18 years or older with a first ICU stay, at least 24 hours of continuous monitoring, and a recorded 12-lead ECG during the index stay were eligible. Patients with a previous admission for the index condition, those discharged within 24 hours, and those with missing key variables were excluded. Of 15,053 heart-failure hospitalizations assessed for eligibility, 376 did not survive the 7-day grace period to time zero and were excluded, leaving 14,677 in the analytic cohort. The deidentified data were approved by the Massachusetts Institute of Technology and Beth Israel Deaconess Medical Center institutional review boards with a waiver of informed consent. The study is reported in accordance with the RECORD-PE statement, the reporting guideline for studies conducted using observational routinely collected health data for pharmacoepidemiology^21^.

#### Target trial protocol

Two target trials were emulated in parallel, with every design decision checked item by item against the TARGET guideline^6^. *Question 1.* Eligible patients were classified into the treatment strategy if they reached at least 50% of the guideline-recommended target dose of an index beta-blocker within 7 days of admission and sustained this exposure through 12 months, and into the control strategy otherwise. *Question 2.* Eligible patients were classified into the treatment strategy if they received GDMT, defined as at least two of the three foundational drug classes (beta-blocker, renin-angiotensin system inhibitor, mineralocorticoid receptor antagonist) at adequate dose for 12 months. Both trials defined time zero as the 24-hour landmark after admission, used intention-to-treat as the primary analysis, and required sustained exposure through 12 months. Per-protocol and as-treated analyses were conducted as sensitivity checks.

#### Estimation

One-year all-cause mortality was the primary outcome. It was estimated by targeted maximum likelihood estimation (TMLE)^2^, a doubly robust, semiparametric efficient estimator. For question 1 (beta-blocker target dose), the treatment mechanism was modeled with a Super Learner^22^ of two candidate learners (main-term logistic regression and Lasso logistic regression), selected by cross-validated out-of-sample risk, and the outcome model with a main-term logistic regression, both fit on the baseline covariates listed in Table 3; for question 2 (guideline-directed medical therapy), the same doubly robust TMLE was used with a main-term logistic regression treatment model (a single Super Learner library, SL.glm), the specification also used for the negative controls. The negative controls were carried on the same hospitalizations over the same one-year window, with the same 7-day grace-period exclusion (Y_W0 = 0), the same 10 baseline covariates, and the same single-point treatment node and estimator as the target estimates; they also share its exposure contrast, because the panel was fitted on the guideline-restricted cohort, in which exposure is defined, as for question 1, by receipt of carvedilol, metoprolol succinate, or bisoprolol at or above 50% of the target dose, which covers 12.1% of the analytic cohort. No element of the negative-control pipeline, including the contrast that defines exposure, therefore differs from the pipeline applied to the target estimates. Of the 12 negative-control outcomes, 10 have uncalibrated relative risks below the null and 2 above it (0.46 to 1.19), so the panel is predominantly but not exclusively protective. They were selected from the OHDSI standard negative-control panel for cardiovascular studies, restricted to outcomes with sufficient event counts and predicted insensitivity to the exposures of interest. The empirical null distribution was jointly estimated as described above. Point estimates were obtained from the bootstrap median of 200 resamples, with credible intervals from the joint Gibbs posterior. Three sensitivity analyses were performed for question 1: leave-one-out across the 12 negative controls; replacement of the Super Learner library; and a Gaussian-process propensity score model with sensitivity parameters drawn from a uniform prior. The emulation is therefore a deliberate best-case stress test: if systematic error survives this combination, it will survive most observational designs.

**Table 2.**
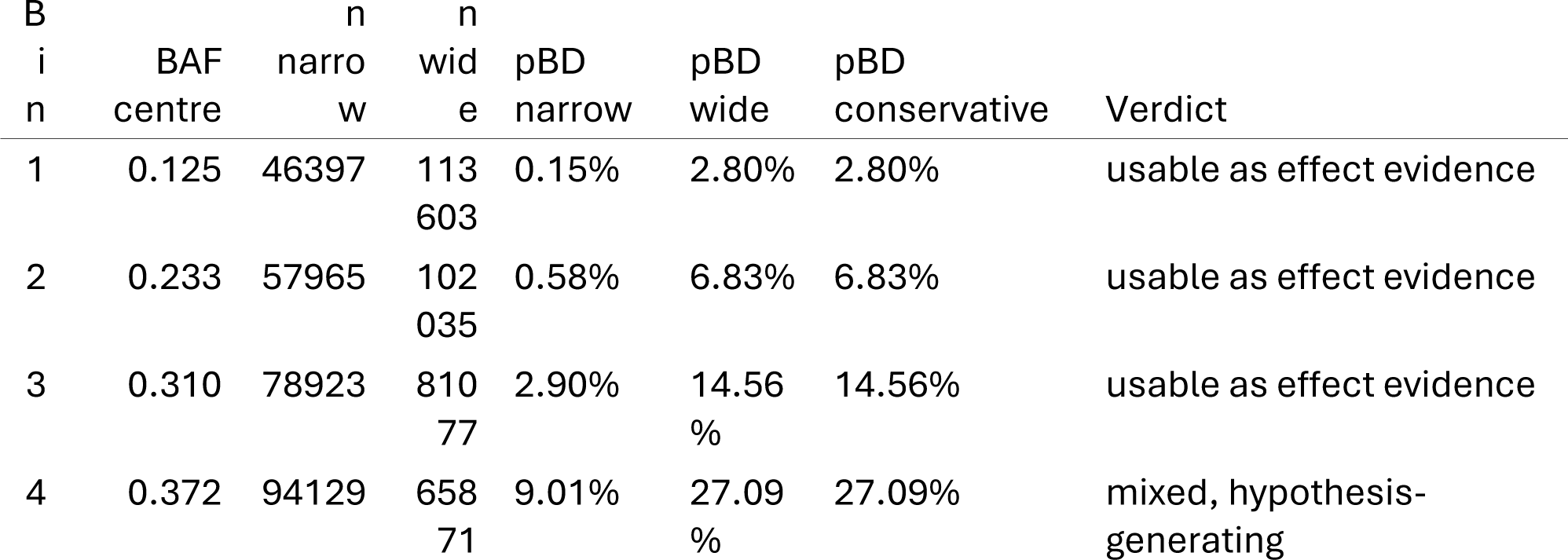
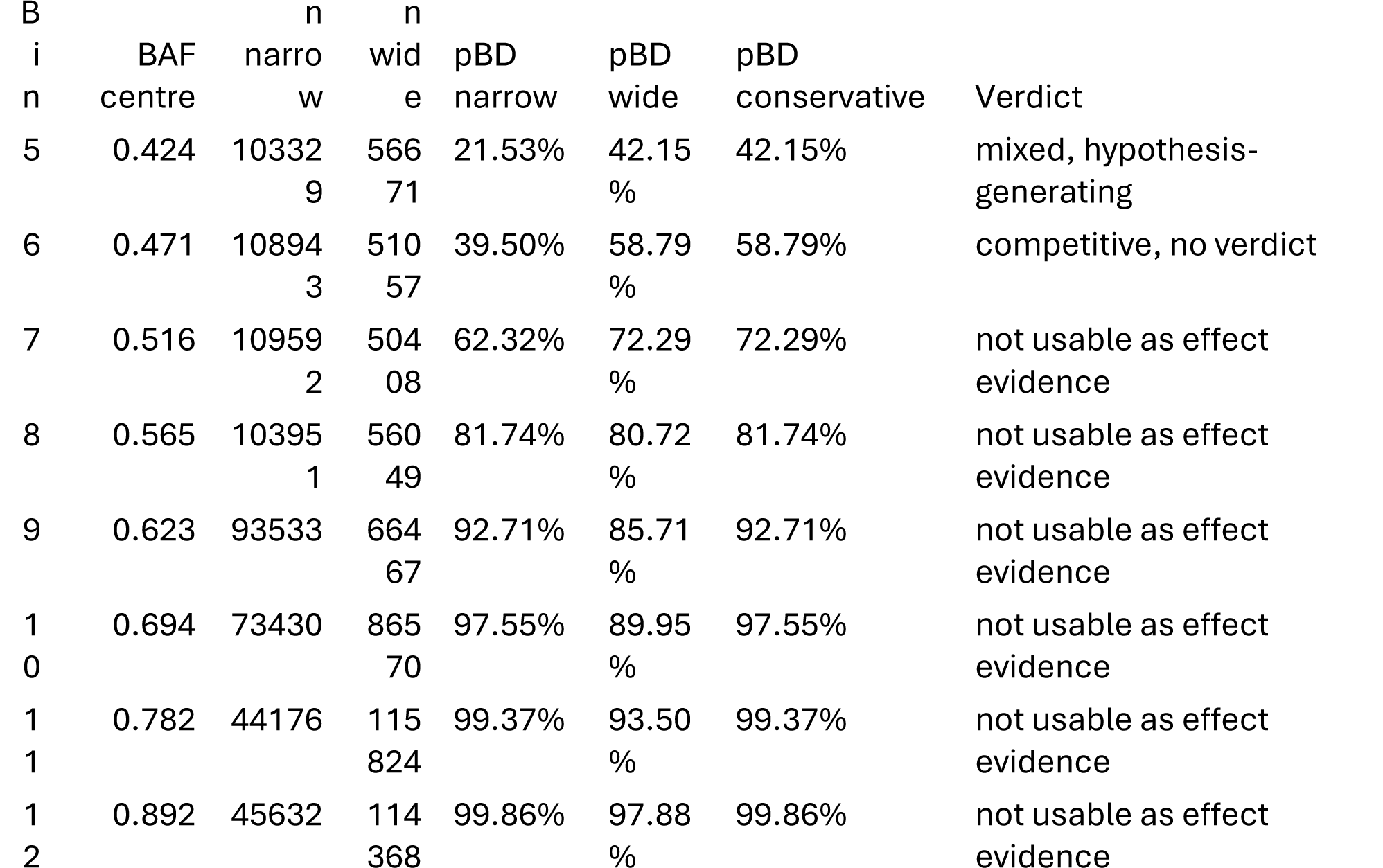
Twelve-bucket equal-frequency lookup table. Each bucket reports the number of repetitions with a narrow and with a wide credible interval, the probability of being bias-dominated under each of those two readings, the conservative reading actually used, and the verdict the conservative reading returns. The credibility intervals are classified as narrow or wide against the median half-width of 0.088 on the BAF scale.

**Table 3.**
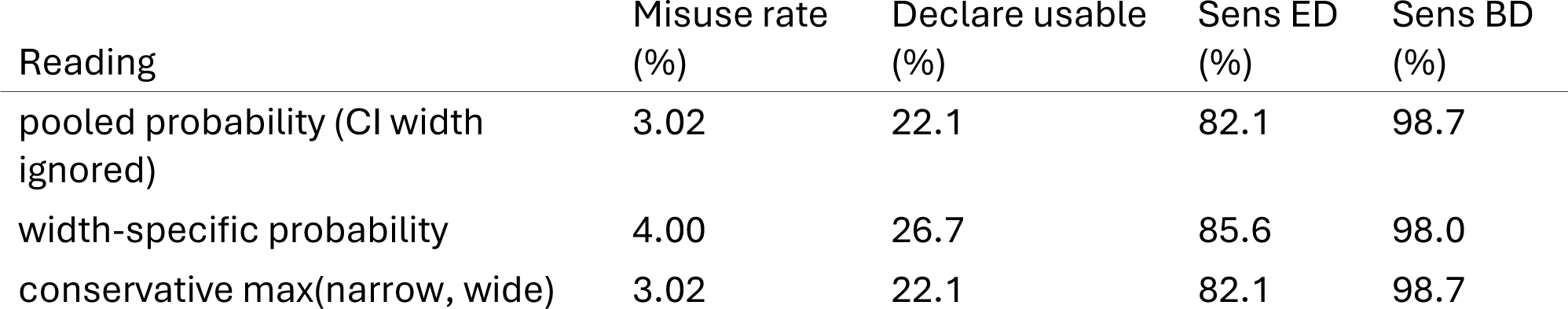
Comparison of three readings of the lookup table: a pooled reading that ignores the credible-interval width, a width-specific reading, and the conservative reading used throughout this manuscript. Sens ED is the share of truly effect-dominated repetitions the reading declares usable; Sens BD is the share of truly bias-dominated repetitions it withholds. The conservative reading changed 0.00 of verdicts relative to the pooled reading, and adding the interval half-width moved the area under the curve by 0.00101.

#### Reporting

For each question we report the uncalibrated relative risk, the calibrated relative risk, the calibrated P-value, BAF̂ with its 95% credible interval, and the verdict from the lookup table. All numbers are reproduced from a single JSON file generated by the analysis pipeline and shared with the companion interactive explainer (https://github.com/zengzhechun/bias-fraction).

## Results

### Part I. Estimator accuracy

Across 1,728,000 interior repetitions, BAF̂ had a mean signed difference (BAF̂ − BAF_*true*_) of -0.020 and a standard deviation of 0.113, yielding 95% limits of agreement of -0.242 to 0.202 (Figure 3). Lin’s concordance correlation coefficient was 0.838. The slope of the regression of the difference on BAF_*true*_ was -0.1003 (95% CI -0.1012 to -0.0995), rejecting the null of no proportional bias. The estimator therefore systematically underestimates BAF_*true*_ and the underestimation grows with the magnitude of BAF_*true*_. Including the *ψ* = -0.01 boundary level widens the limits of agreement to -0.301 to 0.221 and reduces CCC to 0.810. These numbers support an honest reading of the estimator: agreement with truth is moderate, directional underestimation is present, and proportional bias is statistically significant. The estimator is not “in good agreement” with truth in the sense required for direct interpretation. This negative result is the premise for Step 2: a decision rule that uses BAF̂ must be calibrated against truth, and the lookup table in Step 2 does exactly that.

**Figure 1.**
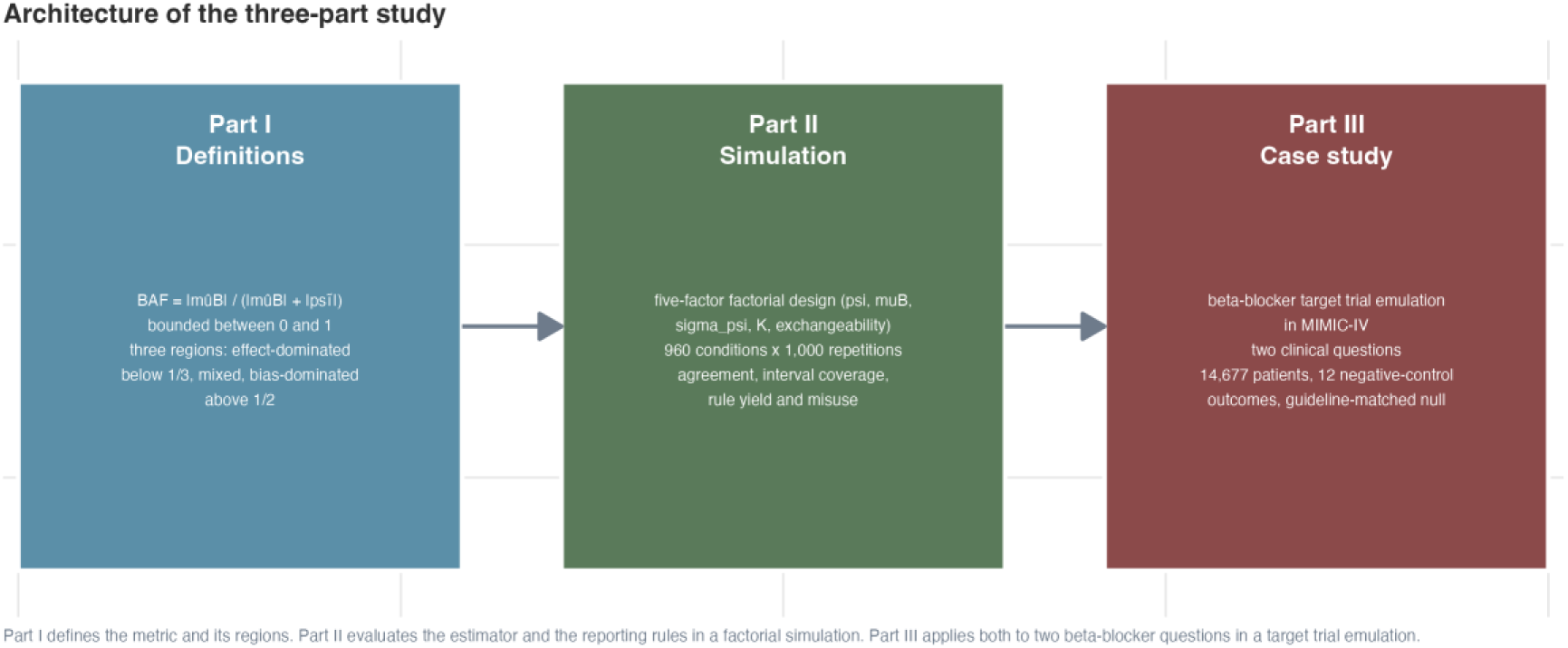
Architecture of the three-part study and the connecting propositions between parts.

**Figure 2.**
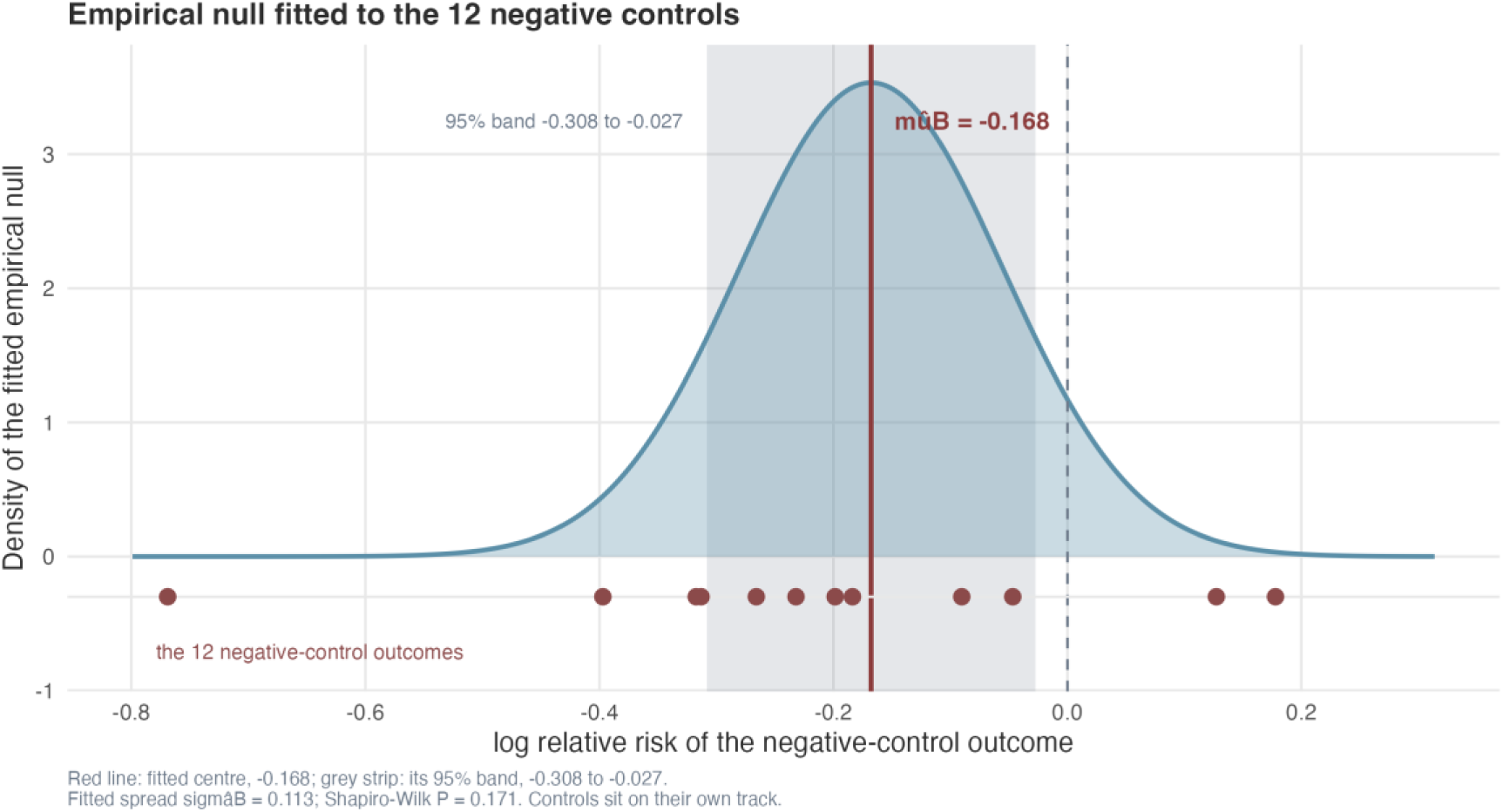
Empirical null distribution from the 12 negative-control outcomes in Part III. The fitted center is marked by the red line; the 12 negative-control observations are shown as filled circles on a separate track to avoid overplotting. *μ̂_B_* = -0.168, *σ̂_B_* = 0.113, Shapiro-Wilk P = 0.171.

**Figure 3.**
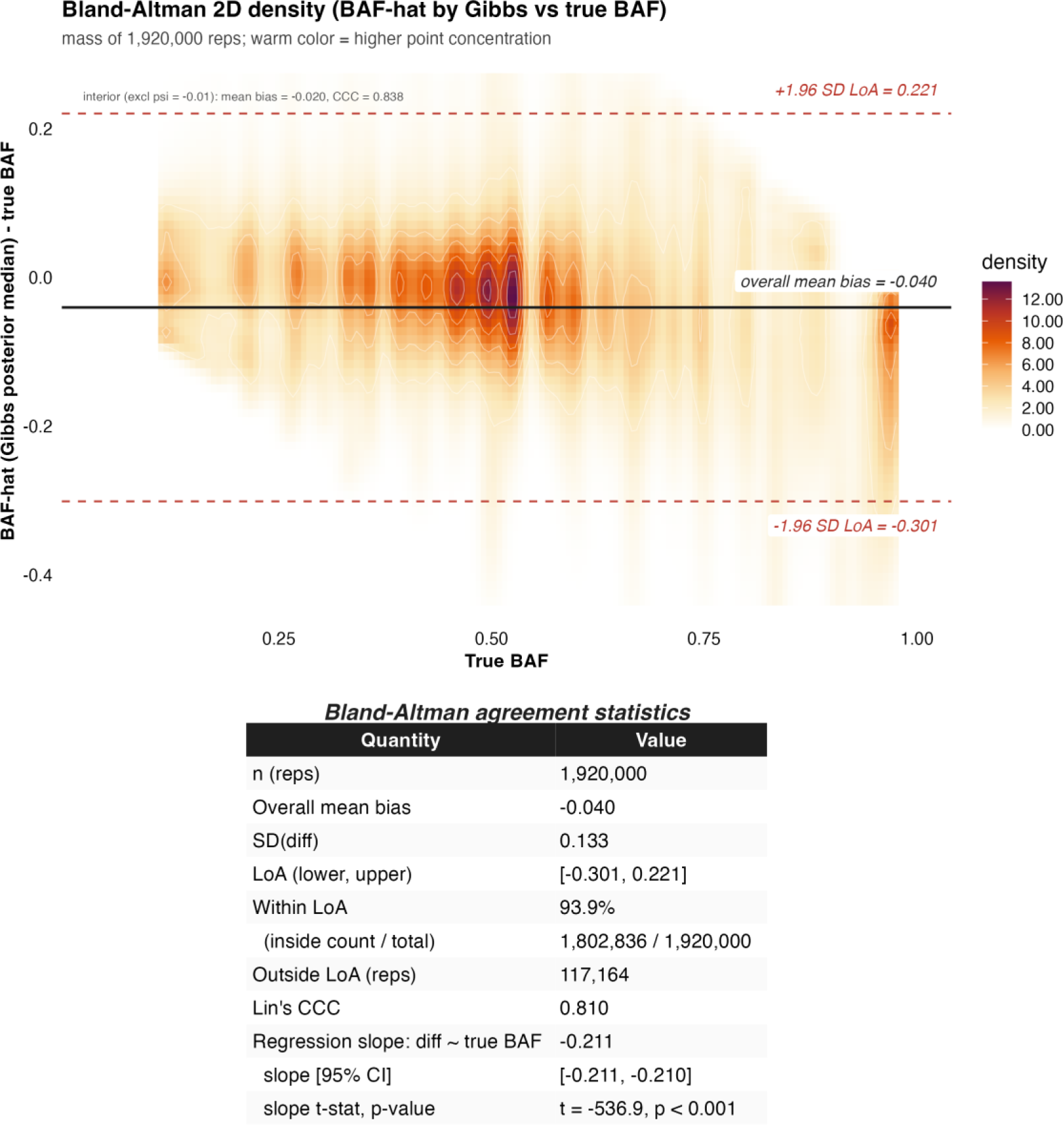
True-BAF-view Bland-Altman agreement of BAF̂, full-grid Monte Carlo simulation (n = 1920000 repetitions across 1920 conditions); the per-repetition empirical null was estimated by Markov chain Monte Carlo. The x-axis is the **true BAF** of the simulated condition; the y-axis is the per-rep signed difference 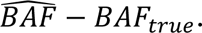 The 2D density (warm ramp on white) summarises all 1,920,000 simulated repetitions at once. The solid horizontal line is the overall mean bias, the two dashed red lines are the 95% limits of agreement (bias ± 1.96 SD), and the inset table reports bias, SD, LoA, Lin’s concordance correlation coefficient, the proportional-bias slope (regression of the difference on *BAF_true_*), and the regression intercept. The estimator systematically underestimates *BAF_true_* (mean bias = -0.040) and the underestimation grows with *BAF_true_* (slope = -0.2106; 95% CI -0.2113 to -0.2098; t = -536.9, P < .001). Lin’s CCC = 0.810 indicates moderate agreement. The interior analysis excluding the *ψ* = -0.01 boundary level (1,728,000 reps) is reported in the supplementary materials.

### Part II. Decision rule performance

#### Layer 1. Calibrated P-value

The calibrated P-value at the conventional 0.05 threshold declared 61.8% of repetitions passable, compared with 67.7% for the uncalibrated P-value. Among those declared passable by layer 1, 36.02% were truly bias-dominated (Table 4). Layer 1 removed one third of the false positives produced by the uncalibrated P-value but still permitted more than one third of the bias-dominated repetitions to pass.

**Table 4.**
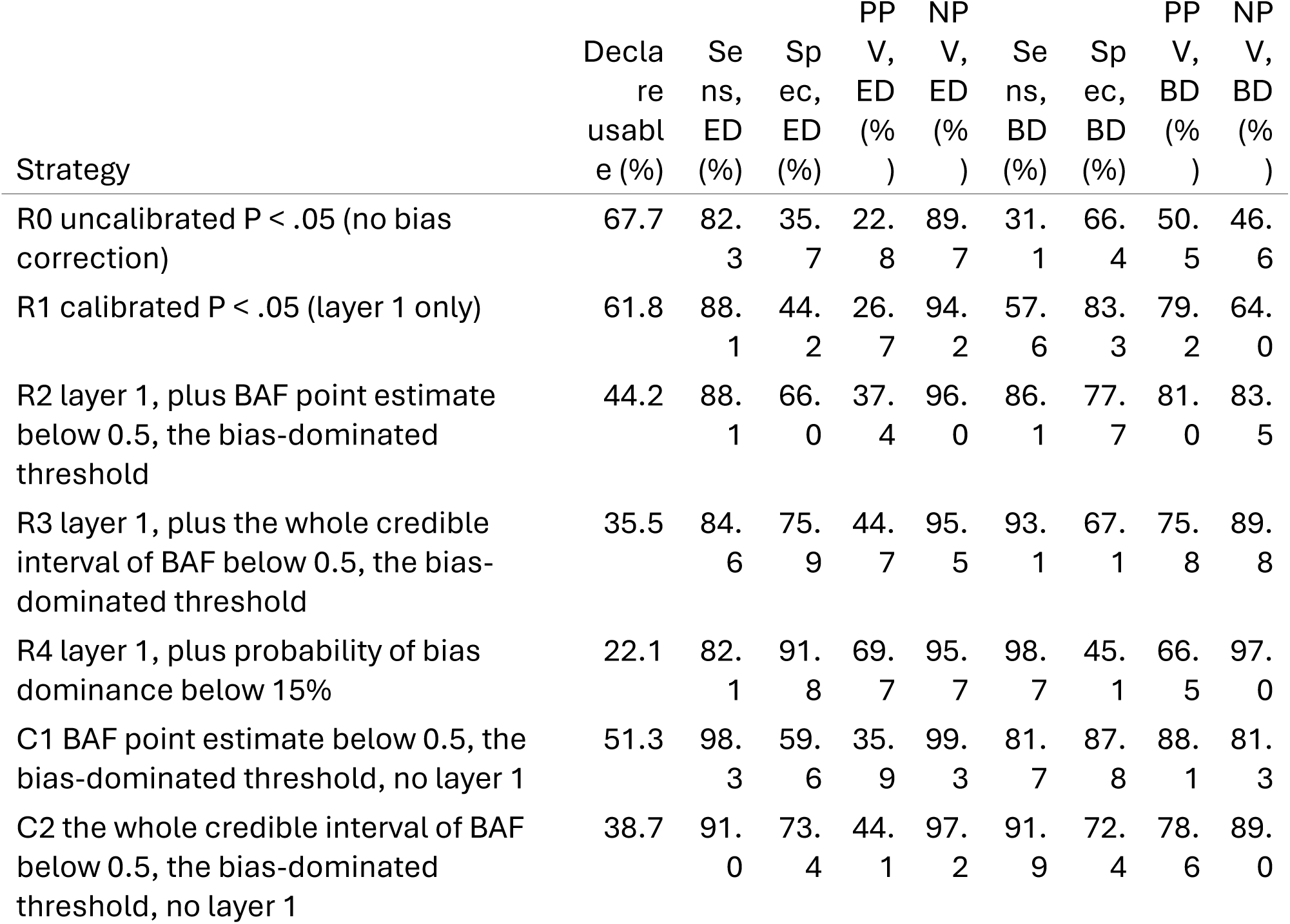
Nested strategy comparison, read from two viewpoints. In the effect-dominated view (ED) a positive call is to declare a repetition usable and a positive truth is to be truly effect-dominated; in the bias-dominated view (BD) a positive call is to withhold and a positive truth is to be truly bias-dominated. The misuse rate is 100 minus NPV in the BD view. Truth has three levels (effect-dominated 18.8%, mixed 28.7%, bias-dominated 52.5%), and the mixed zone is negative in both views, so the views are not mirror images and no pair of columns sums to 100.

#### Layer 2. Calibrated lookup

The lookup table uses BAF̂ and the credible-interval half-width to map each repetition to a probability of being bias-dominated. The continuous relationship between BAF̂ and that probability is monotone, supporting the use of BAF as a credibility scale (Figure 5). Within each of the 12 buckets, the wide intervals and narrow intervals differ in the probability they read by up to 0.206, indicating that interval width carries information. The conservative rule max(*p*_BD, narrow_, *p*_BD,wise_) absorbs this information into the verdict but does not change the verdict relative to the pooled lookup reading; the discriminating action is the rule that the entire 95% credible interval must lie below 0.5, clear of the bias-dominated range.

**Figure 4.**
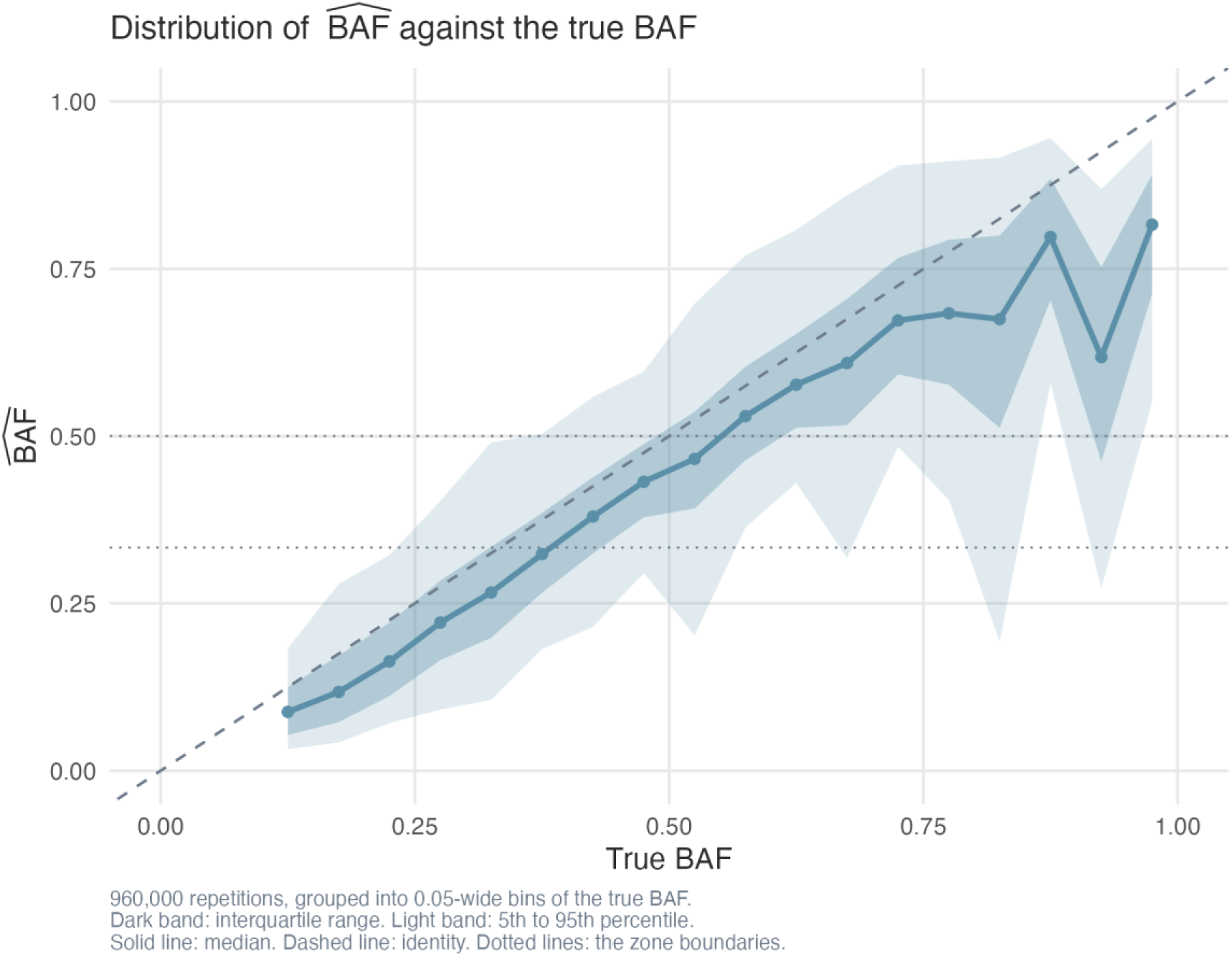
Distribution of BAF̂ against BAF_*true*_. The dark band is the interquartile range, the light band is the 5%-95% range, and the dark blue line is the median. The dashed line is the identity. The estimator systematically underestimates BAF at high BAF_*true*_.

**Figure 5.**
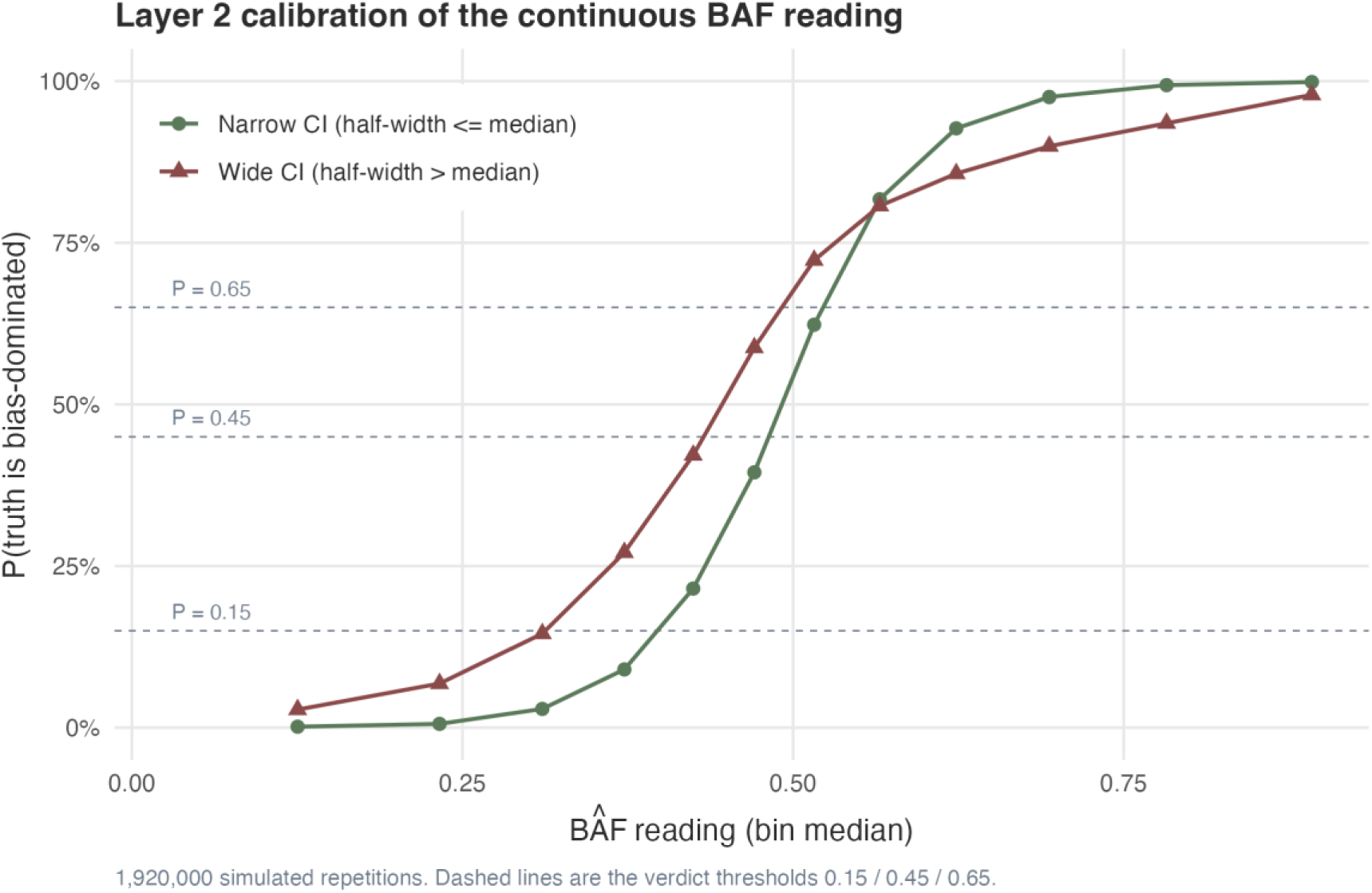
Reliability calibration of the continuous BAF reading. The x-axis is the median BAF̂ within a bucket of the reading; the y-axis is the share of repetitions in that bucket that were truly bias-dominated. Green points are repetitions whose credible interval was at most the median half-width, 0.088, and dark red points those whose interval was wider. Both series are monotone, so BAF behaves as a continuous credibility scale rather than as a yes-or-no test, and a given reading implies a higher probability of bias dominance when the interval is narrow than when it is wide, which is why the interval width is worth reporting next to the point estimate. The dashed lines are the verdict thresholds 0.15, 0.45 and 0.65.

#### Discrimination

Read from the bias-dominated view, meaning that a positive call is to withhold a repetition and a positive truth is to be truly bias-dominated, the area under the ROC curve was 0.787 for the calibrated P-value, 0.922 for BAF̂ alone, and 0.923 for BAF̂ plus interval half-width. Adding the credible interval half-width to BAF̂ yielded a likelihood ratio *χ*^2^ of 3835.1 on 1 degree of freedom (P < .001). The interval half-width was associated with an odds ratio of 3.88 per unit increase. The interval half-width is therefore an informative but minor continuous covariate whose principal role is to enable a defensible interval-level rule, not to improve ranking. The same distinction accounts for the pattern in Table 4: each successive rule improves by imposing a stricter condition, not by ranking repetitions better, which is why the curve in Figure 7 barely moves when the half-width is added. Read from the effect-dominated view, meaning that a positive call is to declare a repetition usable and a positive truth is to be truly effect-dominated, the same three scores reached AUC 0.721 for the calibrated P-value, 0.950 for BAF̂, and 0.950 for BAF̂ plus the half-width, and the half-width score differed from BAF̂ alone by +0.0005, so the sign of the half-width contribution is not consistent across the two views; in neither view does interval width add ranking information beyond BAF̂, and it is retained because rule R3 requires the whole credible interval to fall below 0.5, clear of the bias-dominated zone. BAF̂ therefore ranks the effect-dominated view better (0.95) than the bias-dominated view (0.92), whereas the calibrated P-value does the reverse (0.72 against 0.79). The two scores fail in opposite directions, and only reporting both viewpoints makes that visible. Figure 7 overlays each reporting rule of Table 4 as an operating point, once in each panel, with sensitivity on the vertical axis and one minus specificity on the horizontal axis, so the nested rules can be read directly as the trade-off between catching bias-dominated repetitions and sparing effect-dominated ones.

**Figure 6.**
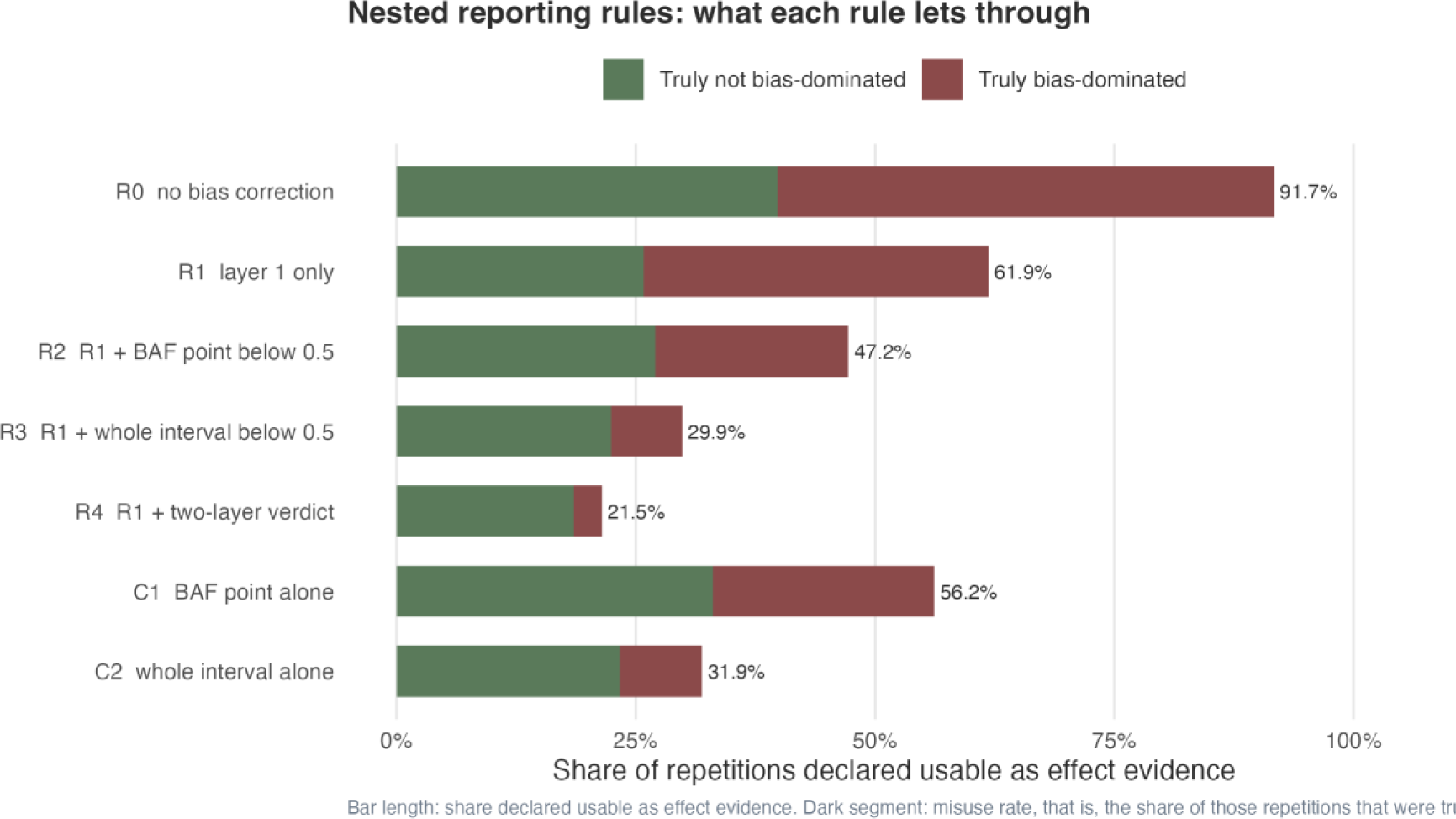
Nested strategy cascade. The bar length is the share of repetitions declared usable by the rule; the dark segment is the share of those repetitions that were truly bias-dominated. Misuse rates decrease from 53.44% (R0) to 3.02% (R4).

**Figure 7.**
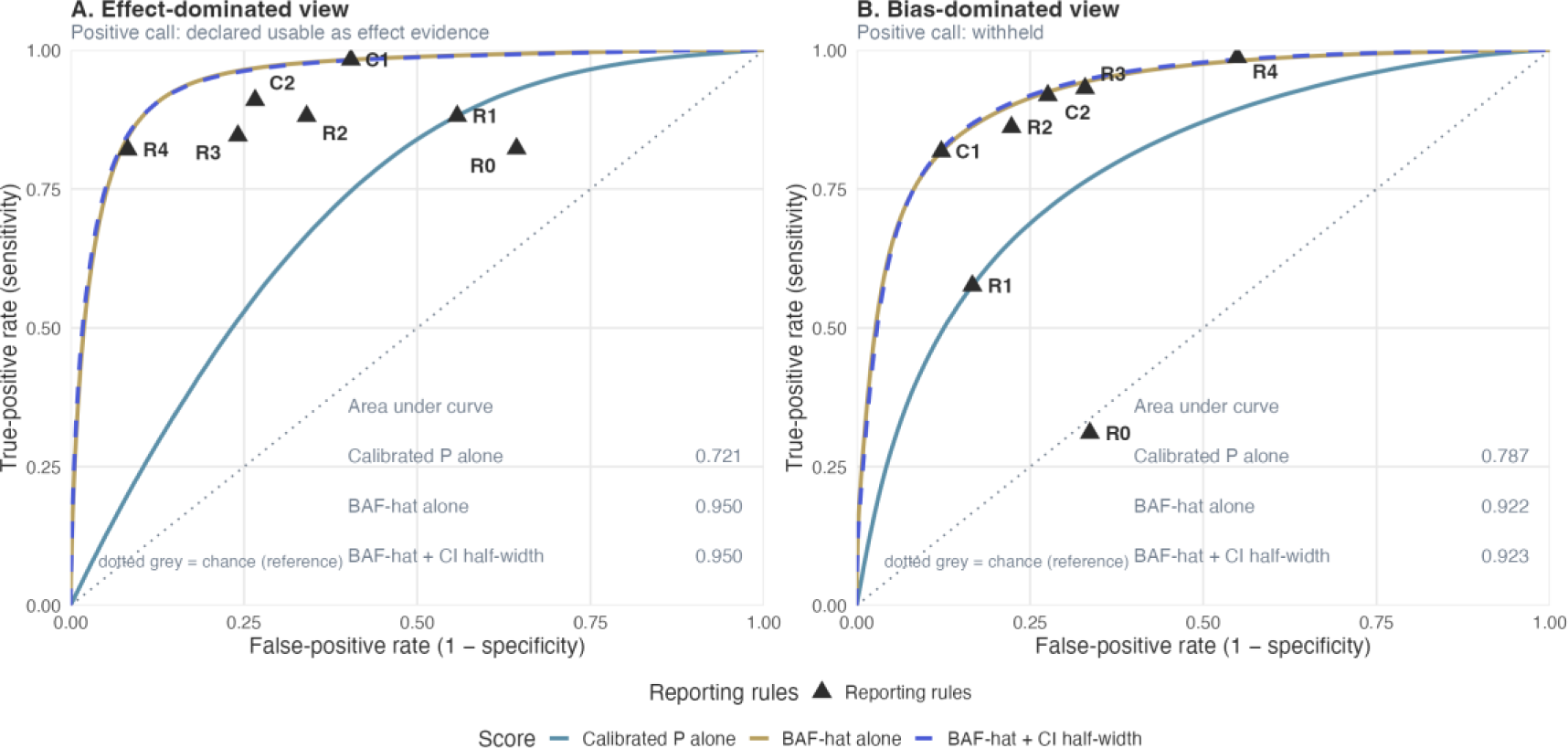
Receiver operating characteristic analysis for the reporting rules of Table 4, read from two viewpoints. Curves: the calibrated P-value alone, BAF̂ alone, and BAF̂ plus its credible-interval half-width; the in-panel tables report the area under each curve in that panel. Triangles: the operating points of R0 through R4 and of C1, C2 of Table 4. Panel A. Effect-dominated view: reference standard truly effect-dominated, positive call declared usable as effect evidence. Panel B. Bias-dominated view: reference standard truly bias-dominated, positive call withheld. Truth has three levels (effect-dominated 18.8%, mixed 28.7%, bias-dominated 52.5%), and the mixed zone falls outside the positive group of both views, which is why the views are not mirror images and why the same curve has a different area in each panel: the calibrated P-value 0.721 in panel A against 0.787 in panel B, BAF̂ 0.950 against 0.922, and BAF̂ plus interval half-width 0.950 against 0.923. All 1,920,000 simulated repetitions (the pooled 1920-condition x 1000-repetition two-grid design) were used for the parametric curve; the simulation itself was not subsampled.

#### Nested strategies

Table 4 reports the misuse rate, the share of repetitions declared usable by the rule that were truly bias-dominated, across nested rules R0 through R4 and stand-alone comparators C1 and C2. The calibrated P-value alone (R1) had a misuse rate of 36.02%. Adding the BAF point estimate below 0.5 (R2) reduced misuse to 16.48%. Adding the credible-interval requirement (R3) reduced misuse to 10.16%. The full rule (R4), meaning a calibrated probability of bias dominance below 15%, reduced misuse to 3.02%, at the cost of declaring 22.1% of repetitions usable (versus 61.8% for R1). Read from the bias-dominated view, meaning that withholding is a positive call and being truly bias-dominated is a positive truth, sensitivity rose from 57.6% for R1 to 98.7% for R4, so the full rule caught a far larger share of the repetitions that truly were bias-dominated, while specificity fell from 83.3% to 45.1%. Read from the effect-dominated view, meaning that declaring usable is a positive call and being truly effect-dominated is a positive truth, sensitivity fell from 88.1% to 82.1% and specificity rose from 44.2% to 91.8%. Truth has three levels and the mixed zone (28.7%) is negative under both views, so the views are not mirror images and no pair of these percentages sums to 100. The stand-alone credible-interval rule C2 achieved a misuse rate of 11.04% without using the calibrated P-value. The BAF estimator alone (C1) achieved a misuse rate of 18.69%.

#### Decision cascade

Figure 9 summarises the rule as applied to 1,920,000 repetitions. Layer 1 passed 1,187,378 (36.0% truly bias-dominated). Within that subset, 424,079 were declared usable as effect evidence (3.02% truly bias-dominated), 272,277 mixed (20.47% truly bias-dominated), 132,047 competitive (45.27% truly bias-dominated), and 358,975 not usable (83.38% truly bias-dominated). The four-zone verdict is monotone in the truth share of bias-dominated repetitions, supporting the use of the lookup as a continuous credibility gauge.

**Figure 8.**
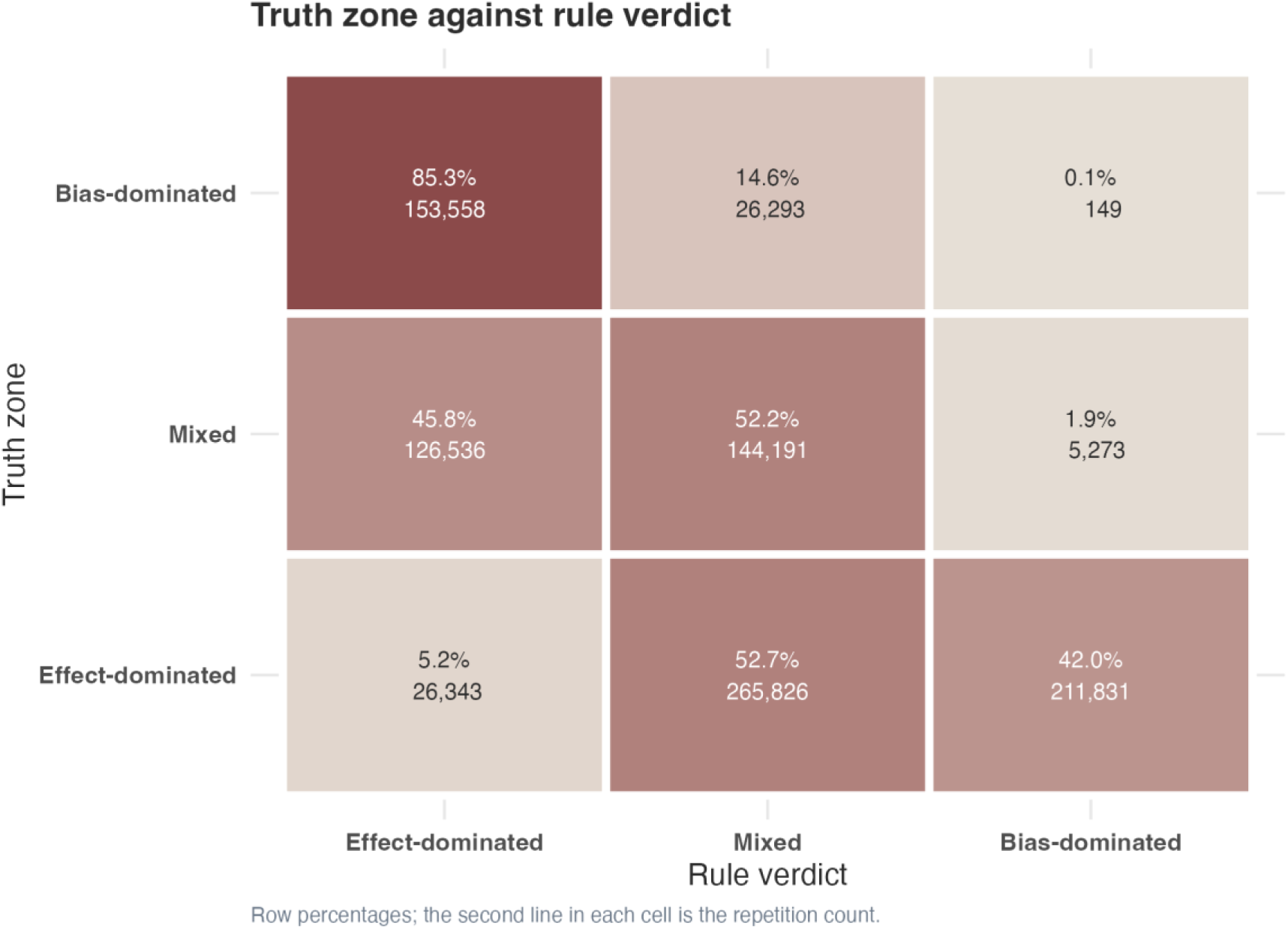
Three-by-three confusion between the truth zone and the verdict returned by the rule, as row percentages. The rule recovers the bias-dominated zone in 85.3% of truly bias-dominated repetitions and the mixed zone in 52.2% of truly mixed repetitions. It almost never returns an effect-dominated verdict: only 5.2% of truly effect-dominated repetitions are called effect-dominated, the rest being returned as mixed (52.7%) or as bias-dominated (42.0%). The asymmetry is consistent with the proportional bias seen in Step 1, and its direction is the conservative one for a rule whose purpose is to withhold an estimate rather than to endorse it.

**Figure 9.**
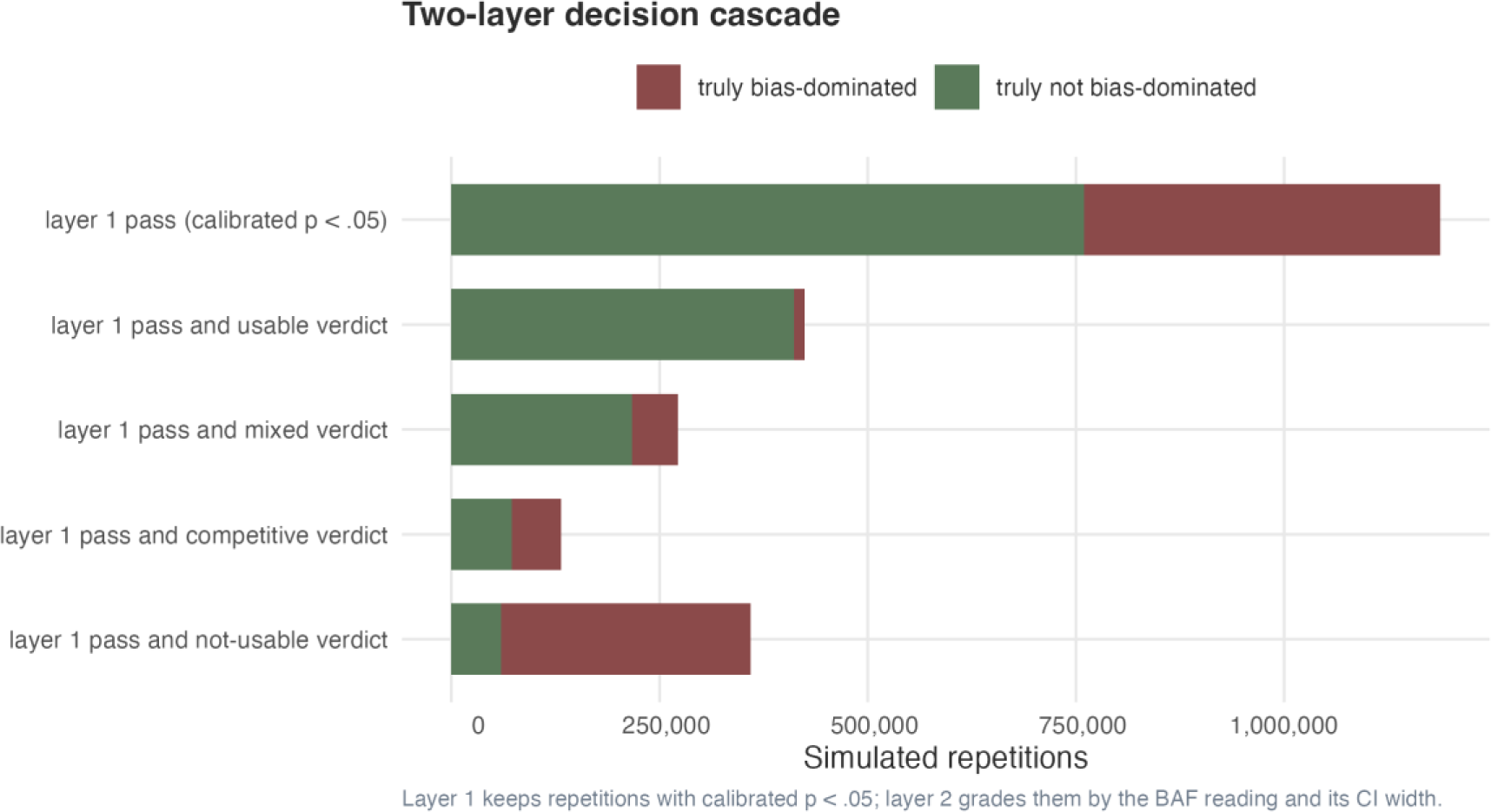
Decision cascade. Each row is one stage of the rule, with the bar length proportional to the number of repetitions reaching that stage and the dark segment proportional to the truth-bias-dominated share within that stage. The four-zone verdict is monotone in the truth share of bias-dominated repetitions.

**Figure 10.**
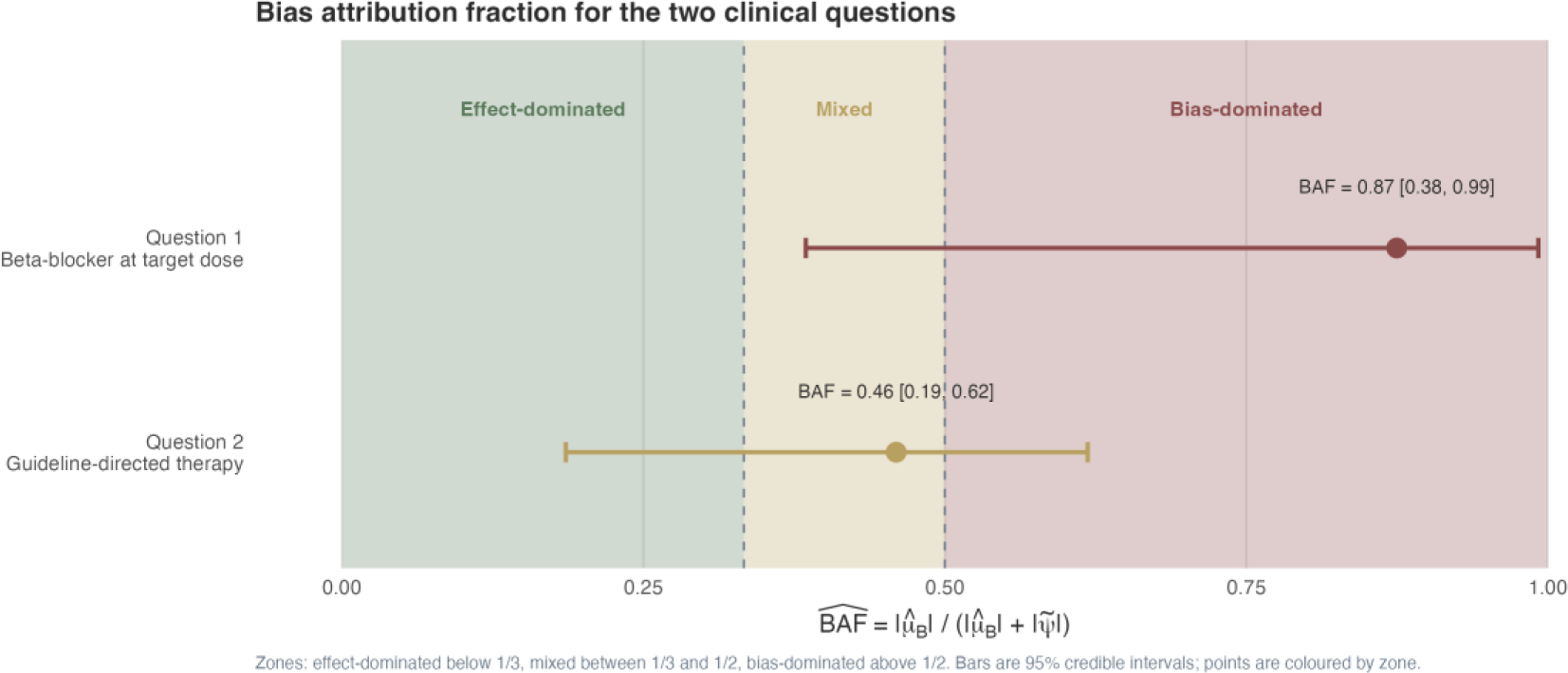
The two clinical questions in Part III plotted on the BAF scale with their 95% credible intervals and verdict zones. Question 1 falls in the “not usable as effect evidence” zone; question 2 falls in the “competitive, no verdict” zone.

#### Robustness

The full rule (R4) was applied to 7 pre-specified subgroups (Table 6). Misuse rates ranged from 1.39% (when bias exchangeability held) to 4.36% (when exchangeability was violated at 30%). The direction of bias in the rule is consistent: exchangeability violation increases misuse. The other 5 subgroups (varying K, σ_*PS*_) gave misuse rates within ±50% of the overall misuse rate, indicating that the rule is not driven by any single design factor.

**Table 5.**
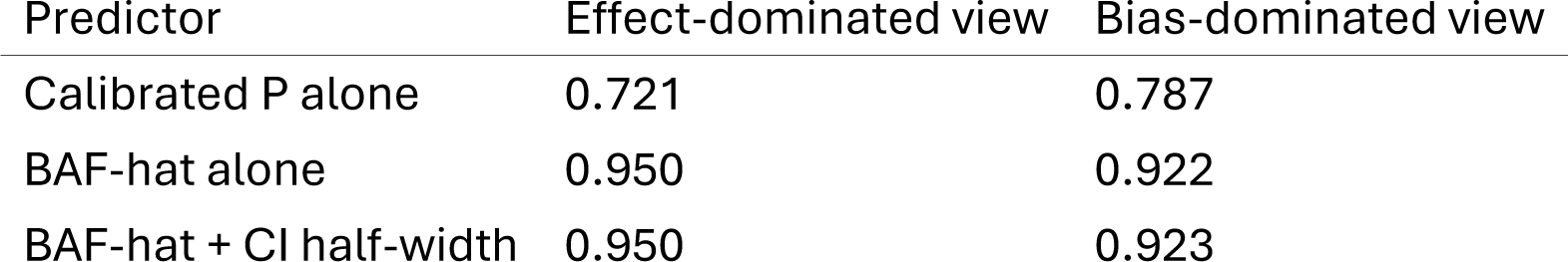
Discrimination of the three continuous readings, from both viewpoints. The area under the curve is reported once for each reading and each view; the same reading therefore has two values, and they are not mirror images. The likelihood-ratio chi-square test for adding the credible-interval half-width to BAFhat is 3835.1 on 1 degree of freedom, P < .001.

| Predictor | Effect-dominated view | Bias-dominated view |
| --- | --- | --- |
| Calibrated P alone | 0.721 | 0.787 |
| BAF-hat alone | 0.950 | 0.922 |
| BAF-hat + CI half-width | 0.950 | 0.923 |

**Table 6.**
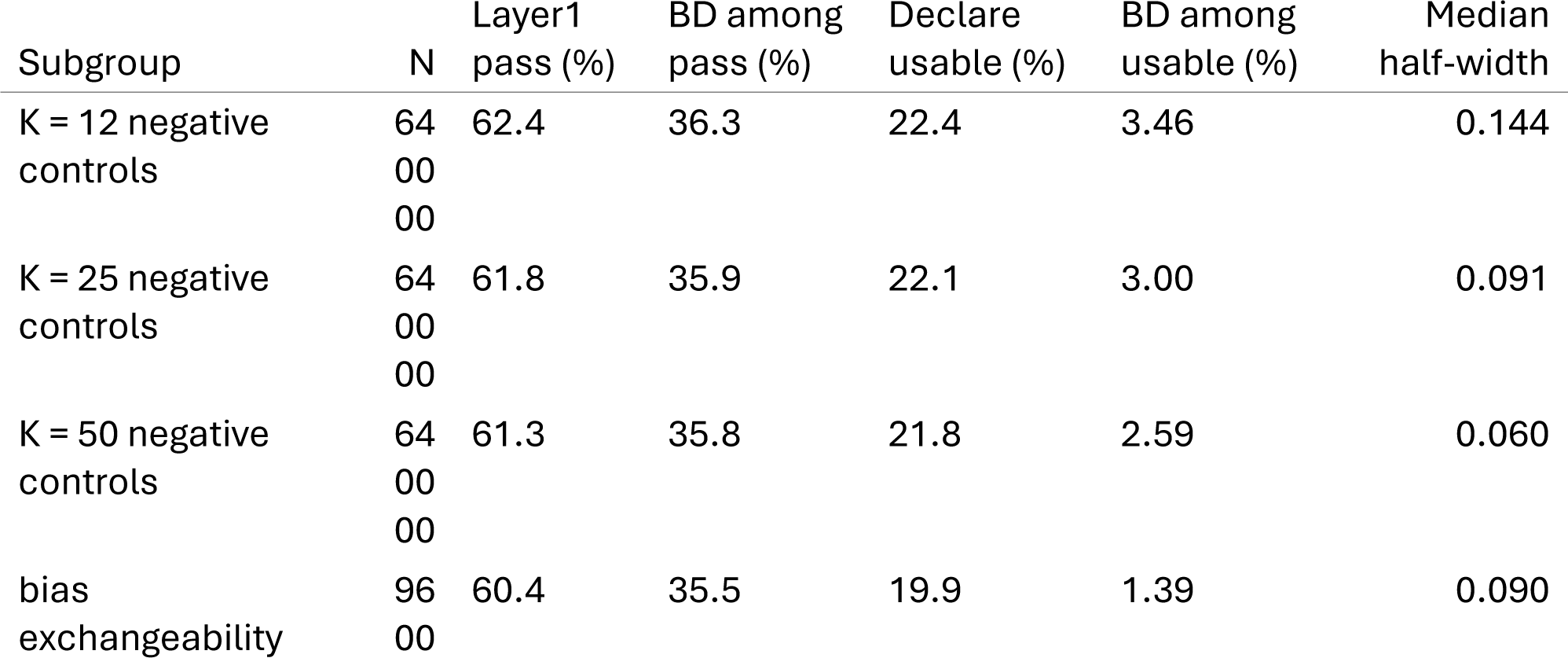
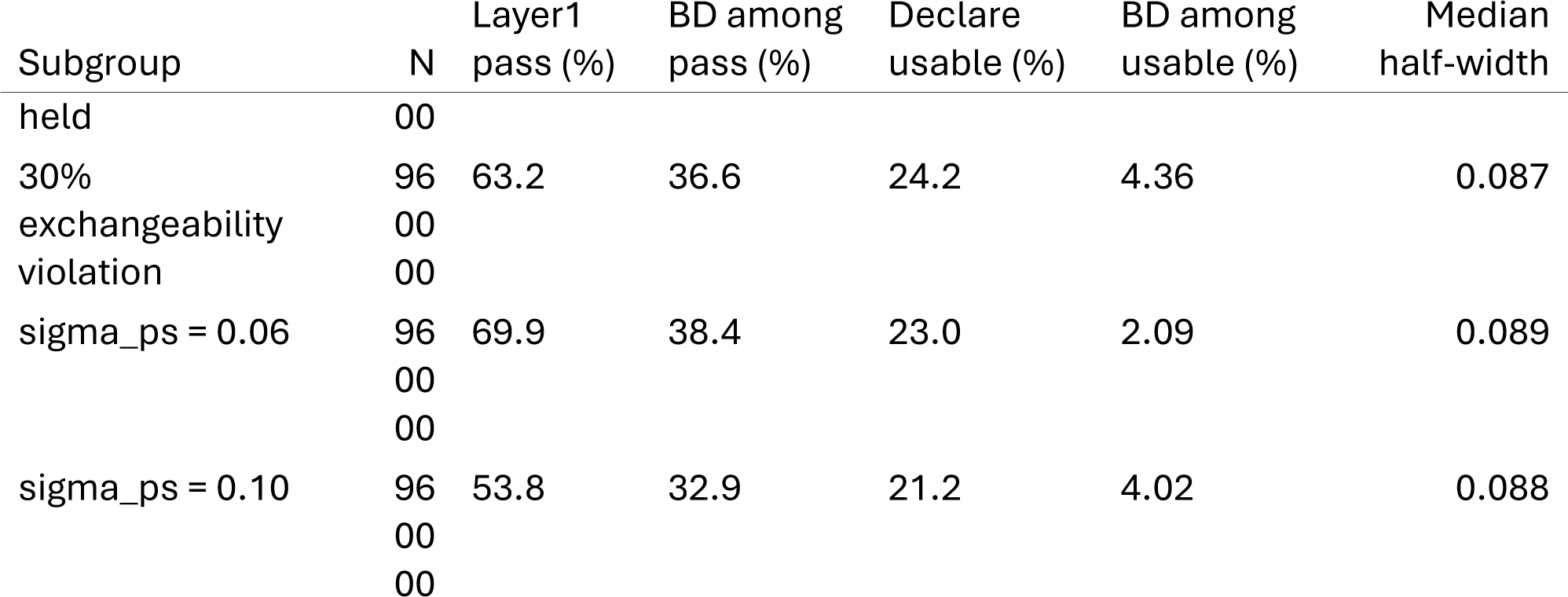
Robustness of the full rule (R4) across the 7 pre-specified subgroups. BD among pass is the share of repetitions the subgroup declares passable at layer 1 that were truly bias-dominated; BD among usable is the misuse rate, that is, the share of repetitions the full rule declares usable that were truly bias-dominated. The subgroup with 30% exchangeability violation produces the largest increase in misuse and is the reason the violation is discussed in Limitations.

### Part III. Beta-blocker target trial emulation

#### Empirical null

Across the 12 negative-control outcomes, the empirical null was estimated as *μ̂_B_* = -0.168, *σ̂_B_* = 0.113. All 12 negative-control estimates fell on the protective side (log relative risks from -0.769 to 0.178), confirming that the systematic bias has a definite direction rather than being random noise. The Shapiro-Wilk P-value of 0.171 does not reject normality.

#### Question 1. Reaching ≥50% of guideline-recommended target dose

The uncalibrated analysis produced a one-year relative risk of 0.831 for the treatment strategy, suggesting a 16.9% reduction in mortality. The calibrated log relative risk was -0.017 and the calibrated relative risk was 0.983. The calibrated P-value was 0.894, well above 0.05, so the layer 1 test did not pass. The signal did not survive the removal of systematic bias. BAF̂ was 0.875 with a 95% credible interval of 0.385 to 0.992 (interval half-width 0.304, classified narrow). The lookup table verdict for this cell was “not usable as effect evidence” with a probability of being bias-dominated of 0.999. Sensitivity analyses left this verdict intact (Table 7). The leave-one-out range of BAF̂ was 0.815 to 0.955; removing the most influential negative-control produced BAF̂ = 0.941. The three alternative treatment-model libraries returned BAF̂ = 0.871, 0.871 and 0.876; two Gaussian-process propensity-score perturbations returned BAF̂ = 0.857 and 0.951.

**Table 7.**
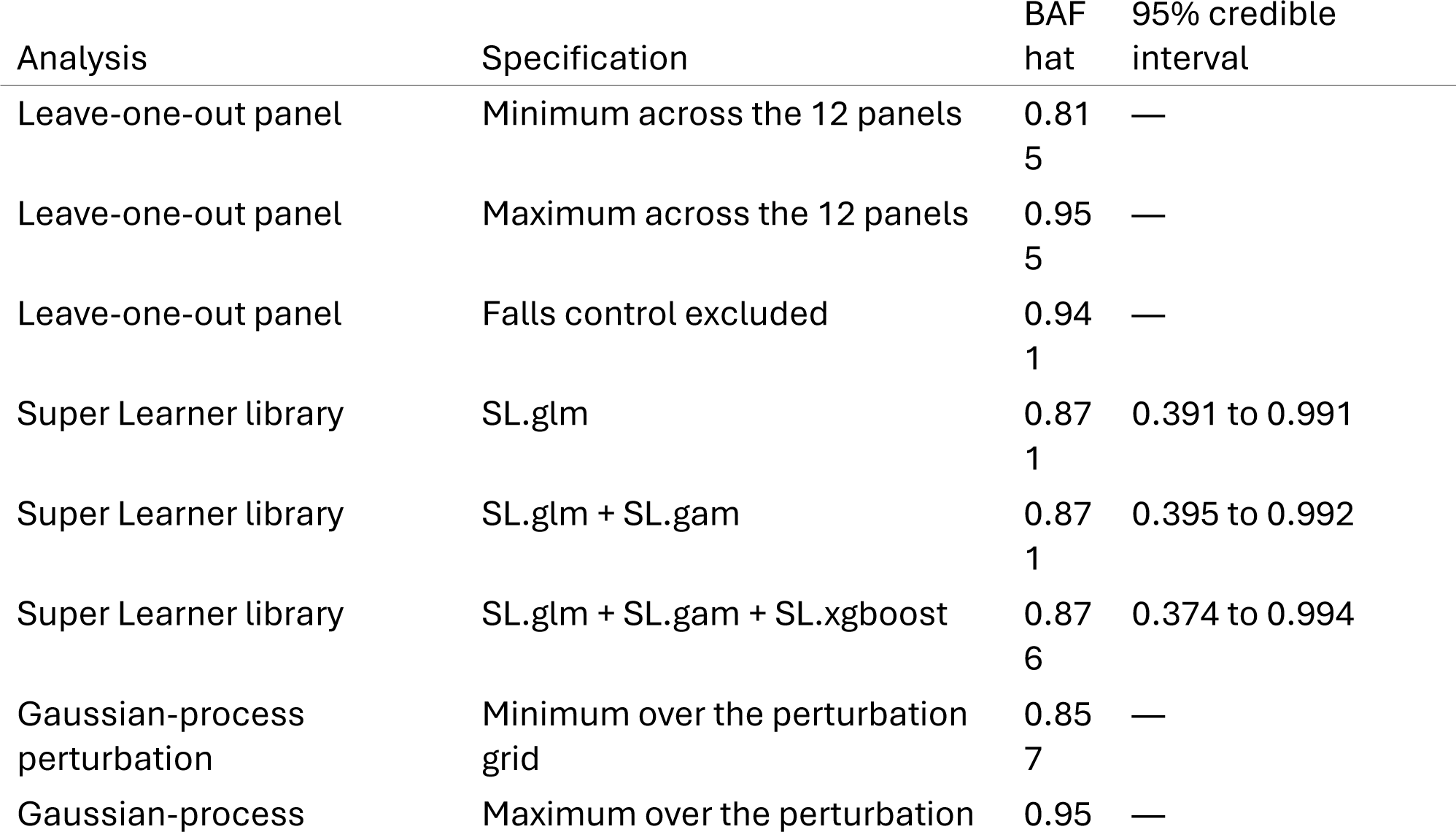
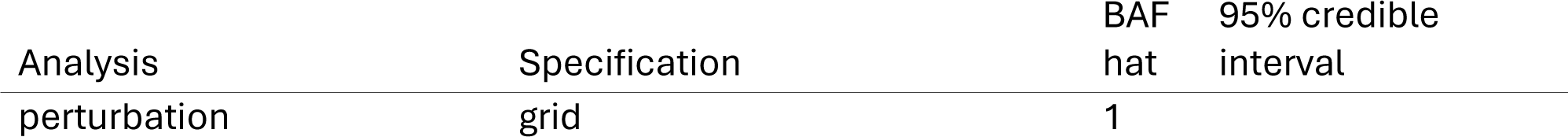
Question-1 sensitivity analyses across three families: leave-one-out resampling of the 12 negative controls, replacement of the Super Learner library, and a Gaussian-process perturbation of the propensity-score model. The credible interval is given for the Super Learner variants; the leave-one-out and Gaussian-process rows report the extremes of a range. The perturbation moved the calibrated relative risk between 0.972 and 1.025 under otherwise identical settings, and every analysis left the question-1 determination at not usable as effect evidence.

| Analysis | Specification | BAF<br>hat | 95% credible<br>interval |
| --- | --- | --- | --- |
| Leave-one-out panel | Minimum across the 12 panels | 0.81<br>5 | — |
| Leave-one-out panel | Maximum across the 12 panels | 0.95<br>5 | — |
| Leave-one-out panel | Falls control excluded | 0.94<br>1 | — |
| Super Learner library | SL.glm | 0.87<br>1 | 0.391 to 0.991 |
| Super Learner library | SL.glm + SL.gam | 0.87<br>1 | 0.395 to 0.992 |
| Super Learner library | SL.glm + SL.gam + SL.xgboost | 0.87<br>6 | 0.374 to 0.994 |
| Gaussian-process<br>perturbation | Minimum over the perturbation<br>grid | 0.85<br>7 | — |
| Gaussian-process<br>perturbation | Maximum over the perturbation<br>grid | 0.95<br>1 | — |

#### Question 2. GDMT with at least two of three classes

The uncalibrated analysis produced a relative risk of 0.682, suggesting a 31.8% reduction in mortality. The calibrated relative risk was 0.807 and the calibrated P-value was 0.1026, above 0.05, so the layer 1 test did not pass. BAF̂ was 0.460 with a 95% credible interval of 0.186 to 0.619 (interval half-width 0.216, classified narrow). The lookup verdict was “competitive, no verdict” with a probability of being bias-dominated of 0.588. The residual association therefore did not survive calibration, and the BAF̂ interval straddled the 0.5 threshold, so the credible evidence cannot distinguish whether what remains is dominated by bias or by effect.

#### Synthesis of the two questions

Question 1 is the headline finding: under the most rigorous design attainable in observational research, an apparent 16.9% mortality reduction attributable to achieving target beta-blocker dose disappears after removing systematic bias. The reported protective association was bias-dominated. None of the safeguards changed this central result: under TARGET-adherent design, doubly robust estimation, and a full negative-control panel, BAF quantified the systematic error that remained. Question 2 confirms that the rule does not over-reject: GDMT has randomized-trial support, and the rule classifies it as a credible signal that nonetheless cannot be reported as effect evidence at the present level of bias control. The two results together demonstrate the three-channel report (signal existence, effect size, and bias fraction) and show that the bias fraction is not negligible even under the strictest design.

## Discussion

This study proposes and evaluates a third reporting channel for observational causal estimates: the fraction of the observed association that is attributable to systematic bias, expressed as a continuous estimator in [0, 1] with a 95% credible interval. The motivation for a third channel is the failure of the binary reporting standards that observational research currently uses. A reported relative risk with a 95% confidence interval answers “how large is the observed effect” but not “how much of the effect is bias.” A reported calibrated P-value answers “is there a signal after removing systematic bias” but not “how much of the remaining signal is still bias.” A bias fraction answers the third question directly.

The principal limitation of the proposal is that BAF inherits the assumption of bias exchangeability from the OHDSI calibration framework. The simulation study showed that a 30% violation of exchangeability raised the misuse rate from 1.39% to 4.36%. This assumption is not testable in any single study and must be justified by domain knowledge about whether the negative controls cover the same bias mechanisms as the target outcome. The interval half-width is informative but is below its nominal 95% coverage in our simulation (0.557 overall, 0.344 narrow, 0.769 wide). The decision rule compensates for this by using a conservative lookup and a separate credible-interval rule, but the rule itself is not a substitute for a calibrated interval.

The estimator has moderate agreement with truth (CCC 0.838) and significant proportional bias, so direct interpretation of BAF̂ is not advised. The lookup table, which is calibrated against truth in a 1,920,000-repetition simulation, absorbs the estimator error. This is a deliberate design choice: the lookup reads as a calibrated probability rather than as a Bayes posterior probability, and the lookup entries are reproducible by any analyst who runs the simulation pipeline.

The contrast between question 1 and question 2 illustrates the operational meaning of the three-channel report. Question 1 fails layer 1: the uncalibrated association did not survive the removal of systematic bias. Reporting this finding as “no association” is consistent with the data and the design. Question 2 fails layer 1 as well, and its BAF̂ interval straddles the 0.5 threshold; the credible evidence cannot distinguish whether the residual association is dominated by bias or by effect. Reporting the pair of questions as “not usable” and “indeterminate” is more honest than reporting either as a positive or as a negative finding.

The reporting protocol proposed here can be applied to any observational study that includes a panel of negative controls, without additional data collection. The empirical null parameters and the calibrated P-value are produced by existing OHDSI tools; the BAF extension and the lookup table are implemented in the companion R package `bafratio`, which retains `bsr_*` names as deprecated aliases. The protocol does not require a validation study of its own; the validation is the simulation study reported here, and the rule parameters are tied to specific simulation conditions that future analysts can replicate or extend.

### Limitations

The empirical null distribution assumes normality; a Shapiro-Wilk test on the present panel did not reject normality, but the rule has not been evaluated on heavy-tailed nulls. The data-generating process in Part II is a simplification: real observational data exhibit time-varying confounding patterns, informative censoring, and outcome-dependent selection that the simulation does not exhaustively represent. The interval half-width information was modelled as a binary split (narrow/wide) rather than a continuous adjustment; a continuous model with a learning-curve adjustment was not significantly better than the binary split in our data. The two clinical questions are illustrative of the method’s behaviour; they are not powered to answer the underlying clinical questions about beta-blocker therapy, which require replication in independent data. Finally, the bias fraction inherits the assumption of bias exchangeability from the calibration framework, which is not testable. Because the primary outcome is all-cause mortality, competing risks do not arise in the primary analysis; they would matter only for cause-specific or nonfatal endpoints, which are used here solely as negative controls. Of the 14,677 patients in the question-2 cohort, 3,494 (23.8%) had a missing guideline-directed medical therapy score and were classified as not optimized; if this missingness is related to the true exposure it is a nonrandom treatment misclassification that biases the question-2 estimate. We therefore re-estimated question 2 under four handling conventions (Table S12): indeterminate treated as suboptimal, as published; complete-case; indeterminate treated as optimized; and multiple imputation from a logistic model of exposure on the baseline covariates. The uncalibrated relative risk moved from 0.68 to 0.91 across conventions, but no convention produced a calibratable signal, and none yielded a usable determination: the bias fraction rose from 0.460 under the published convention to between 0.463 and 0.676 under the others, and 2 of the four conventions placed question 2 in the least favourable determination of not usable as effect evidence. The published convention is thus the most favourable of the four, so the qualitative conclusion does not depend on it. The negative-control panel was refitted on the guideline-restricted cohort so that it shares the exposure contrast of the target estimates, and every result reported here uses that panel. Realigning it widened the empirical null (SD 0.068 under the earlier all-agent panel versus 0.113 here) and moved the question-2 calibrated P value from 0.045 to 0.103, so the layer-1 pass question 2 would have obtained under the earlier panel was not robust to this realignment: with the realigned panel neither question passes layer 1, while both determinations are unchanged, question 1 remaining not usable as effect evidence (calibrated P 0.894 versus 0.946 before realignment) and question 2 remaining a competitive determination with no usable finding.

As defined in the Methods, the interval is a credible interval, not a confidence interval: it propagates draws of *μ̂_B_* while holding the observed effect estimate fixed, so it carries no frequentist coverage guarantee. In the merged simulation pool it contained BAF_*true*_ in 55.7% of repetitions (71.1% in the primary grid alone, where bias and effect share a sign), below the nominal 95%. Two features of the construction contribute to this gap. First, the interval omits the sampling variability of the effect estimate. Second, the number of draws is modest (M = 300 in the simulation; a bootstrap of comparable size in the case study), so the 2.5th and 97.5th percentiles carry Monte Carlo uncertainty. Propagating the omitted term jointly restores coverage: adding the sampling variability of the calibrated estimate raised coverage from 78.8% to 97.7% across the conditions examined, at the cost of widening the median half-width from 0.132 to 0.222 and lowering the yield of the strictest interval rule from 24.3% to 13.2% of repetitions (Table S11). Because the interval is systematically narrower than its nominal coverage, the rules that consume its upper endpoint are affected, and not in the direction usually assumed. R3, and its case-study analogue C2, declare an estimate usable only when the entire interval lies below 0.5, a condition a narrower interval satisfies more easily; the same narrowing also makes the interval more often classified as narrow by the width split of the lookup table. Restoring nominal coverage therefore lowers the yield of these rules rather than raising it, and because the repetitions that drop out are those with the least evidence against bias dominance, the misuse rate falls with it: propagating the omitted term jointly lowered the yield of R3 from 24.3% to 13.2% of repetitions and its misuse rate from 5.1% to 0.6% (Table S11). The R3 misuse rate reported here is therefore an upper bound on the rate that would obtain at nominal coverage, not a lower bound.

The lookup table that maps 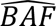 and interval half-width to a determination was built from the same 1,920,000 repetitions used to evaluate the rules, so its reported performance reflects in-sample agreement and is optimistically biased. Two held-out validations bound that optimism (Table S10). Holding out 20% of repetitions within every condition left every rule’s misuse rate unchanged to two decimal places. Holding out 20% of the 1920 conditions instead, which also tests grid cells that never contributed to the lookup, left the share declared usable essentially unchanged and raised the misuse rate of the strictest rule from 2.7% to 2.8%, the largest excursion across the five folds being 4.0%. Because the data-generating truth is fixed by the investigator, absolute misuse rates are illustrative and depend on the chosen ranges of the simulation factors; what is robust is the relative ordering of strategies, which held across the prespecified subgroups, including the exchangeability-violation level. The direction of the bias is a further restriction: the case study belongs to the regime in which bias inflates the apparent effect. The performance numbers reported here pool the two half-grids and therefore average over all four roles; the companion grid with the sign flipped (Table S15 and Table S16) shows that the point estimate stays accurate when the bias shrinks, cancels, or reverses an effect, whereas the credible interval becomes markedly narrower and its coverage falls further, and that the three-region classification is more stable to the direction of the bias than the interval is.

### Conclusions

Reporting the calibrated P-value alongside BAF̂ and its 95% credible interval provides a three-channel report of signal, effect size, and bias fraction. In a 1,920,000-repetition simulation, the three-channel report reduced the rate at which bias-dominated repetitions were declared usable as effect evidence from 53.44% to 3.02%. In a target trial emulation of two beta-blocker questions in MIMIC IV, the three-channel report classified question 1 as not usable as effect evidence and question 2 as competitive, no verdict. The protocol can be applied to any observational study that uses negative controls.

## Data Availability

The MIMIC-IV database (version 2.2) is available through PhysioNet (https://physionet.org/content/mimiciv/2.2/) and the MIMIC-IV-ECG extension (version 1.0) at https://physionet.org/content/mimic-iv-ecg/1.0/ to credentialed users under the PhysioNet data use agreement. All analysis code, including the simulation pipeline, the case-study TMLE estimation pipeline, and the BAF computation, is available at https://github.com/zengzhechun/bias-fraction. The methods are implemented in the companion bafratio R package (version 0.4.0) at https://github.com/zengzhechun/bafratio.

---

Table 1. Five-factor simulation design. The five factors (true effect ψ, bias center μ_*B*_, standard error of the target estimate σ_*ψ*_, number of negative controls K, exchangeability violation) were crossed in a full factorial design within each of two half-grids that differ only in the sign of the bias center, yielding 1920 conditions in the pooled design.

Table 2. Twelve-bucket lookup table. Each bucket reports the count of repetitions with narrow and wide credible intervals and the conservative probability of being bias-dominated. Verdict zones are colour-coded.

Table 3. Comparison of three readings of the lookup (pooled, width-specific, conservative). The conservative reading is the one used in the manuscript.

Table 4. Nested strategy comparison, read from two viewpoints. Every rule withholds a repetition unless all the conditions listed for it are met; only then is the repetition declared usable as effect evidence. Misuse rate is the share of repetitions declared usable by the rule that were truly bias-dominated, which is 100 minus NPV in the bias-dominated view. A single rule has two readings at once. The effect-dominated view (ED) takes a positive call to be “declared usable” and a positive truth to be truly effect-dominated; the bias-dominated view (BD) takes a positive call to be “withheld” and a positive truth to be truly bias-dominated. In each view, Sens is the share of truly positive repetitions the rule called positive, Spec the share of truly negative repetitions the rule called negative, PPV the share of repetitions the rule called positive that were truly positive, and NPV the share it called negative that were truly negative. Truth has three levels (effect-dominated 18.8%, mixed 28.7%, bias-dominated 52.5%), and the mixed zone is negative in both views, so the views are not mirror images and no pair of columns in this table sums to 100. C1 and C2 are stand-alone comparators that do not use the calibrated P-value. The operating point of each rule is plotted in Figure 7, once in each panel.

Table 5. Discrimination, read from two viewpoints. In the bias-dominated view a positive call is to withhold a repetition and a positive truth is to be truly bias-dominated; in the effect-dominated view a positive call is to declare a repetition usable and a positive truth is to be truly effect-dominated. The calibrated P-value reached an area under the ROC curve of 0.787 (bias-dominated view) and 0.721 (effect-dominated view); BAF̂ reached 0.922 and 0.950; and BAF̂ plus credible-interval half-width reached 0.923 and 0.950. The likelihood-ratio χ² test for adding credible-interval half-width to BAF̂ is 3835.1 on 1 degree of freedom (P < .001).

Table 6. Robustness. Misuse rate across 7 pre-specified subgroups. Exchangeability violation at 30% is the most important subgroup and produces the largest increase in misuse.

Table 7. Question 1 sensitivity analyses. Leave-one-out across the 12 negative controls; replacement of the Super Learner library; Gaussian-process propensity-score perturbations.

## Declarations

### Ethics approval and consent to participate

The deidentified data were approved by the Massachusetts Institute of Technology and Beth Israel Deaconess Medical Center institutional review boards with a waiver of informed consent, as stated in the Methods.

### Author Contributions

Dr Zeng had full access to all of the data in the study and takes responsibility for the integrity of the data and the accuracy of the data analysis. Dr Zeng conceived the study, designed the credibility metric, implemented the analysis code and the bafratio R package, conducted the simulation study, and drafted the manuscript. Dr Wang contributed to the methodological design and reviewed the statistical approach. Dr Zuo contributed to the clinical interpretation of the negative control selection and reviewed the manuscript. Dr Shu contributed to the study design and critically revised the manuscript for important intellectual content. All authors read and approved the final manuscript. Dr Zeng is the author and maintainer of the bafratio R package used in this work.

### Data Sharing Statement

The MIMIC IV database (version 2.2) is available through PhysioNet (https://physionet.org/content/mimiciv/2.2/) and the MIMIC-IV-ECG extension (version 1.0) at https://physionet.org/content/mimic-iv-ecg/1.0/ to credentialed users. All analysis code, including the simulation pipeline, the case-study TMLE estimation pipeline, and the BAF computation proposed in this work, is available at https://github.com/zengzhechun/bias-fraction, together with an interactive graphical explainer of the metric, the simulation, and the two clinical questions. The methods proposed here are implemented in the companion bafratio R package (version 0.4.0; https://github.com/zengzhechun/bafratio), released under the MIT license, which provides the BAF estimator with bootstrap and Fieller intervals, the two-layer screening rule, the full set of reporting rules evaluated in Table 2, leave-one-out sensitivity analysis, and negative control diagnostics, together with a unit-test suite covering the estimator, the interval procedures, and the classification thresholds. The EmpiricalCalibration R package (Schuemie et al.), which supplies the empirical null machinery used here but does not include BAF, is available from CRAN.

### Additional Contributions

The authors thank the MIMIC IV team for making the database publicly available and the OHDSI/EmpiricalCalibration R package developers for the empirical calibration tools upon which this work builds.

### Artificial Intelligence Use Disclosure

The authors used artificial intelligence tools (DeepSeek V4 Pro, Kimi K3, GLM 5.3, ChatGPT 5.5, and Tencent WorkBuddy) for code development, manuscript editing, and language polishing. All AI-assisted content was critically reviewed and revised by the authors, who take full responsibility for the content of this manuscript.

## Supplementary appendix

The supplementary appendix (separate document) reports additional simulation details, the per-lookup-bucket sensitivity analysis, and the full case-study estimation output.

