## Supplementary Appendix for "Quantifying Systematic Bias in Observational Causal Estimates Using Negative Controls: The Bias Attribution Fraction"

Zeng Zhechun

Wang Jinwen

Zuo Huijuan

Shu Lixia

2026-08-27

^1^ Beijing Anzhen Hospital, Capital Medical University, Beijing 100029, China

^2^ Beijing Institute of Heart, Lung and Blood Vessel Diseases, Beijing, China

This supplementary appendix provides the methodological detail, full lookup table, and complete case-study estimation output that support the main manuscript. All numbers are reproduced from the analysis outputs of the study repository (https://github.com/zengzhechun/bias-fraction).

### S1. Extended methods

#### S1.1 Full simulation design

The simulation crossed five factors in a full factorial design within each of two half-grids that differ only in the sign of the bias center: the primary grid, and a companion grid of identical size generated with the same procedure and a disjoint random-number stream (seed 100,043 versus 43), which flips the sign of μ_B at every level while holding all other factors fixed. The pooled design comprises 1920 conditions, each replicated 1000 times for a total of 1,920,000 repetitions, and every performance analysis in the main manuscript and in this appendix is computed on that pooled evidence base. The factors and levels were:

- True effect on the log relative risk scale, ψ, with 10 levels from -0.40 to -0.01.
- Systematic bias center, μ_B, with 8 magnitudes given at both signs (negative in the primary grid, positive in the companion grid), from −0.40 to −0.05 and +0.05 to +0.40 in steps of 0.05.
- Standard error of the target estimate, σψ, with 2 levels (0.06, 0.10).
- Number of negative controls, K, with 3 levels (12, 25, 50).
- Exchangeability violation, with 2 levels (0, 0.3, where 0.3 denotes a 30% violation rate).

The data-generating process was a logistic regression in which each negative-control estimate was drawn as Y_k ~ N(μ_B, σ_B^2 + s_k^2), with the target estimate generated under the analogous model. By construction, each repetition carried a known truth BAF_{true} in [0, 1], an estimated BAF̂, and a calibrated P-value. The design allocated 864,000 interior repetitions (excluding the ψ = -0.01 boundary level) and 96,000 boundary repetitions. The truth composition was 52.5% bias-dominated, 28.7% mixed, and 18.8% effect-dominated.

#### S1.2 Bias Attribution Fraction definition and interval construction

The Bias Attribution Fraction is BAF = |μ̂_B| / (|μ̂_B| + |ψ̃|), where μ̂_B is the estimated center of the empirical null and ψ̃ is the calibrated log relative risk. Because BAF is a ratio of two jointly estimated quantities, the delta method is unstable when ψ̃ is near zero. The credible interval was therefore constructed by propagating uncertainty rather than approximating it: a set of draws of μ_B was generated from the fitted systematic error model, and each draw was passed through the same function BAF = |μ_B| / (|μ_B| + |Y - μ_B|), with the observed target estimate Y held at its observed value. The 2.5% and 97.5% empirical quantiles of the resulting BAF distribution formed the interval, and the point estimate was the median of that same distribution, so an estimate always lies inside the interval reported for it. In the simulation the draws were posterior draws from the systematic error model Y_k ~ N(μ_B, σ_B^2 + s_k^2), fitted by a Gibbs sampler with priors μ_B ~ N(0, 1) and σ_B^2 ~ InverseGamma(0.5, 0.05); in the case study they were nonparametric bootstrap resamples of the negative-control panel, with the empirical null refitted within each resample. Both constructions yield a distribution for BAF itself, and a bootstrap percentile distribution can be read as an approximation to a posterior under an uninformative prior, so a single term, credible interval, is used for both.

#### S1.3 Negative-control selection for the case study

The negative-control panel was assembled before any case-study estimate was produced, and no control was added or removed afterwards. A candidate was an outcome with no plausible causal pathway from either treatment strategy and therefore expected to show a null association with it. Screening proceeded in four levels, summarised in Table S13.

The candidate list contained 20 outcomes chosen for biological plausibility of a null association and for coverage of distinct clinical domains, spanning infection, gastrointestinal, trauma, ophthalmology, rheumatology, urology, oncology, dental, and vascular disease.

Four candidates were removed on code feasibility. Fracture, burn, and poisoning are recorded with external-cause codes, and inguinal hernia is recorded surgically; neither family is captured in the MIMIC-IV diagnoses table, and each had no records there. That left 16 candidates with usable code definitions.

The remaining candidates had to reach at least 50 hospitalizations in the heart-failure cohort, counted as distinct patients carrying the code in the index hospitalization or in any later hospitalization. 2 candidates failed this level, Conjunctivitis and Non-Melanoma Skin Cancer, with 44 and 38 qualifying patients respectively, leaving 14.

Finally the event count had to reach 50 inside the analysis window itself, which excludes patients who died during the 7-day grace period and counts only events occurring after it. 2 further candidates failed here, Dental Caries and Varicose Veins, with 45 and 36 events in that window, leaving the 12 controls of the reported panel.

Both quantitative thresholds were fixed before the panel was screened and were applied mechanically, so the panel size was determined by the data source rather than by the results. No candidate was removed on the basis of the strength or the direction of its estimated association, and the panel is not the subset of candidates that happened to appear null: 2 of the 12 retained controls in fact have uncalibrated relative risks above 1. The reported panel is therefore predominantly but not exclusively protective, with calibrated log relative risks ranging from -0.769 to 0.178 (10 below the null and 2 above it). A Shapiro-Wilk test of the empirical null residuals yielded p = 0.171, consistent with approximate normality of the null.

#### S1.6 Heart failure cohort and the ejection-fraction proxy

Eligibility was anchored on a heart-failure discharge diagnosis rather than on measured left ventricular function. Heart failure was defined as any ICD-10-CM diagnosis code beginning with I50 or any of the ICD-9-CM codes 428.0, 428.1, 428.20 through 428.23, 428.30 through 428.33, 428.40 through 428.43, and 428.9. Because neither MIMIC-IV nor MIMIC-IV-ECG records left ventricular ejection fraction, no ejection-fraction threshold could be applied, and the cohort mixes reduced with preserved systolic function. The proxy used to test whether that heterogeneity drives the case-study result is defined in S5.5.

#### S1.7 Beta-blocker exposure and guideline target dose

For question 1 the exposure was receipt, during the 7 days after the first electrocardiogram, of a beta-blocker with Class I evidence in heart failure at or above 50% of its guideline target dose. Three agents qualified, with these target daily doses: carvedilol 50 mg, metoprolol succinate 200 mg, and bisoprolol 10 mg. Beta-blockers without Class I evidence in this population, namely metoprolol tartrate, atenolol, nebivolol, propranolol, and sotalol, did not count toward exposure, so a patient receiving only one of them is classified as unexposed.

The daily dose was reconstructed from the hospital prescription record rather than taken from a study-specific instrument. The recorded dose value was multiplied by the recorded number of doses per 24 hours, with a missing or non-positive value of the latter set to once daily; the product was expressed as a percentage of the drug’s target dose, computed separately for each agent; and percentages above 200% were set missing, because a value that extreme reflects a recording unit rather than a plausible dose. Dose was then summarised per patient as the maximum percentage across qualifying prescriptions whose start date fell in the window. A patient was coded as at or above the target dose (level 2) when that maximum reached 50%, as partially exposed (level 1) when a qualifying beta-blocker was recorded below 50%, and as unexposed (level 0) when no qualifying prescription was found. In the analytic cohort the distribution was 10,621 (72.4%) unexposed, 2,284 (15.6%) partially exposed, and 1,772 (12.1%) at or above the target dose; question 1 compares the last group with the two others combined.

Question 2 used the same dose reconstruction within each of three foundational drug classes rather than for a single agent. The classes were renin-angiotensin system inhibitors (angiotensin-converting enzyme inhibitors, angiotensin receptor blockers, and the angiotensin receptor-neprilysin inhibitor), mineralocorticoid receptor antagonists, and beta-blockers. A class counted toward the score when at least one agent within it reached 50% of its target dose, and the score was the number of classes that counted, so that the exposed group for question 2 was a score of at least 2 of 3. Target daily doses were as given above for beta-blockers and, for the other two classes, were lisinopril 35, enalapril 20, ramipril 10, captopril 150, quinapril 40, trandolapril 4, fosinopril 40, benazepril 40, perindopril 8, moexipril 30, valsartan 320, losartan 150, candesartan 32, irbesartan 300, olmesartan 40, telmisartan 80, and sacubitril with valsartan 400 mg/day for renin-angiotensin system inhibitors, and spironolactone 50 and eplerenone 50 mg/day for mineralocorticoid receptor antagonists.

#### S1.8 Exposure definition shared by the negative-control panel and the target estimates

The negative-control panel and the target estimates are produced on the same cohort, the same 10 baseline covariates, the same 7-day grace period, and the same estimators; the only quantity that differs is the outcome. In particular the empirical null that calibrates both case-study estimates is fitted on the same guideline-restricted exposure contrast that defines the target estimates, rather than on a contrast that counts all beta-blocker agents. Misalignment on this point is not innocuous: under the broader all-agent contrast the fitted between-control standard deviation was 0.068 and the question-2 calibrated P value 0.045, whereas under the aligned contrast they are 0.113 and 0.103; the wider null and the larger calibrated P value are the values used throughout this manuscript.

#### S1.4 Construction and calibration of the BAF credible interval

BAF is a ratio whose denominator is itself the difference between two estimated quantities, so its uncertainty cannot be inherited from a standard error in the usual way. As ψ̃ approaches 0 the ratio becomes unstable, and the instability runs in one direction: the denominator shrinks, so BAF is pushed toward 1, while the upper end of the BAF scale is a hard boundary at exactly 1. An interval built on a normal approximation for BAF̂ would be symmetric on a scale that is bounded and asymmetric near its end, and would put mass outside [0, 1]. We therefore propagate uncertainty instead of approximating it, as described in S1.2.

Holding the observed effect estimate fixed is a deliberate choice: the interval answers how much of the calibrated signal is bias given the effect estimate that was obtained, not a joint statement about the effect estimate and the empirical null. In the simulation the number of retained joint posterior draws was modest (M = 300, from 400 Gibbs iterations with 100 discarded as warm-up), which is too few to locate the 2.5th and 97.5th percentiles precisely and is therefore itself a source of interval instability.

We call the result a credible interval, not a confidence interval, because it is a percentile range of a distribution for the quantity being estimated, not the output of a procedure constructed to cover the true value in 95% of repeated samples. No sampling distribution is inverted, so the interval carries no guarantee of frequentist coverage. The distinction is not terminological: in the simulation, where BAF_true is known by construction, the interval contained it in 55.7% of repetitions, below the nominal 95%, and coverage was lower among narrower intervals (34.4%) than among wider ones (76.9%). One reason is visible in the construction itself: the interval omits the sampling variability of the effect estimate, a second source of error in BAF̂. Reported as a credible interval with its coverage stated, this is an honest statement that the propagated distribution is too narrow rather than a failure of the method.

Each interval is summarised by its half-width, (BAF_hi − BAF_lo)/2, and not by its two bounds. BAF is bounded, so the bounds are truncated at 0 and 1 and carry information about location as well as precision; the half-width extracts the precision component in BAF units, on the same bounded scale as the estimate itself, which is what lets the lookup table of Step 2 classify an estimate jointly by where it falls on the BAF scale and by how wide its interval is, splitting the half-width at its median across the simulation (0.088) and returning the higher of the two stratum-specific probabilities. The half-width is not a standard error: because the bounds are truncated, it is half of the [0, 1] scale that the interval occupies, and near either end of the scale it is attenuated by that truncation.

#### S1.5 Justification of the simulation factor ranges

Factor levels were not chosen to represent a survey of cardiovascular observational research. Each level was anchored either to a quantity measured in the present case study or to a published estimate of adherence-related bias, as follows.

| Factor | Levels | Values | Justification | Source |
| --- | --- | --- | --- | --- |
| True effect, ψ | 10 | −0.01 to −0.40 | Spans marginal to large protective effects on the log relative-risk scale (RR, 0.99 to 0.67) and brackets the beta-blocker estimate observed in the case study (ln RR, −0.18) | Present case study |
| Bias center, μB | 8 | −0.05 to −0.40 | (RR, 0.95 to 0.67). Approaches, without reaching, the healthy-adherer effect reported for adherence-related exposures (odds ratio, 0.56; ln, −0.58) and brackets the empirical null fitted to the case-study panel (μ̂B, −0.17) | Simpson 2006; present case study |
| Standard error of the target estimate, σψ | 2 | 0.06, 0.10 | Closely matches the standard errors of the two case-study estimates (0.056 and 0.068), the smaller falling marginally below the lower grid level | Present case study |
| Number of negative controls, K | 3 | 12, 25, 50 | 12 is the number of controls in the cardiovascular panel available in MIMIC-IV; 25 is a moderately larger panel; 50 tests whether estimator performance keeps improving or plateaus once the panel far exceeds the size typically assembled in practice | Present case study; Schuemie 2018 |
| Exchangeability violation | 2 | 0, 0.3 | 0 corresponds to an assumption that holds; 0.3 shifts 30% of controls by +0.15 (RR, 1.16), a deliberately arbitrary perturbation chosen to break the assumption rather than to reproduce any empirical estimate | Design choice |

Three respects in which the grid does not bracket the case study should be read as limitations of the simulation, all of which make it more favorable to the estimator than the case study itself. First, the between-control standard deviation was held fixed at 0.05, below the 0.113 fitted to the case-study panel. Second, control standard errors were drawn from a uniform distribution on 0.03 to 0.12, whereas the observed controls ranged from 0.025 to 0.302 with 3 controls above 0.24. Third, the guideline-directed medical therapy estimate (ln RR, −0.38) lies between grid levels, within the ψ grid whose most extreme value is −0.40.

### S2. Part I supplementary: Bland-Altman agreement

The interior analysis (primary) excludes the ψ = -0.01 boundary level to avoid near-boundary pathologies. The full analysis includes it. Both are reported below.

Table S1. Bland-Altman agreement of BAFhat as an estimator of BAFtrue.

| Analysis | N | Mean.diff..BAFhat.BAFtrue. | SD.diff | LoA.low | LoA.high | CCC | Prop..bias.slope | MAE | RMSE |
| --- | --- | --- | --- | --- | --- | --- | --- | --- | --- |
| Interior (primary) | 1728000 | -0.02 | 0.113 | -0.242 | 0.202 | 0.838 | -0.1003 | 0.085 | 0.115 |
| Full (incl. boundary) | 1920000 | -0.04 | 0.133 | -0.301 | 0.221 | 0.810 | -0.2106 | 0.099 | 0.139 |

The worst single-condition mean signed difference among the 1728 interior conditions was 0.321. The proportional-bias slope regression rejected the null of no proportional bias in both analyses (interior slope -0.1003, 95% CI -0.1012 to -0.0995; full slope -0.2106, 95% CI -0.2113 to -0.2098). The estimator systematically underestimates BAF_{true}, and the underestimation grows with the magnitude of BAF_{true}.

### S3. Part II supplementary: decision-rule detail

#### S3.1 Full 12-bucket lookup table

Each bucket reports the count of repetitions with narrow and wide credible intervals, the probability of being bias-dominated under the narrow reading, the wide reading, and the conservative max(narrow, wide) reading, together with the verdict zone.

Table S2. Twelve-bucket equal-frequency lookup table mapping BAFhat x interval width to probability of being bias-dominated.

| Bin | BAF.centre | n.narrow | n.wide | pBD.narrow | pBD.wide | pBD.cons. | Verdict |
| --- | --- | --- | --- | --- | --- | --- | --- |
| 1 | 0.125 | 46397 | 113603 | 0.15% | 2.80% | 2.80% | usable as effect evidence |
| 2 | 0.233 | 57965 | 102035 | 0.58% | 6.83% | 6.83% | usable as effect evidence |
| 3 | 0.310 | 78923 | 81077 | 2.90% | 14.56% | 14.56% | usable as effect evidence |
| 4 | 0.372 | 94129 | 65871 | 9.01% | 27.09% | 27.09% | mixed, hypothesis-generating |
| 5 | 0.424 | 103329 | 56671 | 21.53% | 42.15% | 42.15% | mixed, hypothesis-generating |
| 6 | 0.471 | 108943 | 51057 | 39.50% | 58.79% | 58.79% | competitive, no verdict |
| 7 | 0.516 | 109592 | 50408 | 62.32% | 72.29% | 72.29% | not usable as effect evidence |
| 8 | 0.565 | 103951 | 56049 | 81.74% | 80.72% | 81.74% | not usable as effect evidence |
| 9 | 0.623 | 93533 | 66467 | 92.71% | 85.71% | 92.71% | not usable as effect evidence |
| 10 | 0.694 | 73430 | 86570 | 97.55% | 89.95% | 97.55% | not usable as effect evidence |
| 11 | 0.782 | 44176 | 115824 | 99.37% | 93.50% | 99.37% | not usable as effect evidence |
| 12 | 0.892 | 45632 | 114368 | 99.86% | 97.88% | 99.86% | not usable as effect evidence |

#### S3.2 Three readings of the lookup

The lookup supports a pooled reading (CI width ignored), a width-specific reading, and the conservative max(narrow, wide) reading used in the manuscript.

Table S3. Comparison of three readings of the lookup table. Sens ED, share of truly effect-dominated repetitions declared usable; Sens BD, share of truly bias-dominated repetitions withheld.

| Reading | Misuse.rate…. | Declare.usable…. | Sens.ED…. | Sens.BD…. |
| --- | --- | --- | --- | --- |
| pooled probability (CI width ignored) | 3.02 | 22.1 | 82.1 | 98.7 |
| width-specific probability | 4.00 | 26.7 | 85.6 | 98.0 |
| conservative max(narrow, wide) | 3.02 | 22.1 | 82.1 | 98.7 |

The conservative reading changed 16.28% of verdicts relative to the pooled reading and 0.00% relative to the width-specific reading; the maximum absolute within-bin probability gap was 0.206, and adding CI half-width moved the AUC by only 0.00101.

#### S3.3 Nested strategy comparison

Strategies R0 through R4 add one component at a time; C1 and C2 are stand-alone comparators that do not use the calibrated P-value. Every rule is scored twice, once from each viewpoint of Table 4 in the main text, so Table S4 reports eight operating characteristics per rule. The two readings are not mirror images because truth has three levels and the mixed zone is negative in both.

Table S4. Nested strategy comparison, read from two viewpoints. In the effect-dominated view (ED) a positive call is to declare a repetition usable and a positive truth is to be truly effect-dominated; in the bias-dominated view (BD) a positive call is to withhold and a positive truth is to be truly bias-dominated. Sens, share of truly positive repetitions the rule called positive; Spec, share of truly negative repetitions the rule called negative; PPV, share of repetitions the rule called positive that were truly positive; NPV, share the rule called negative that were truly negative. The misuse rate reported in Table 4 of the main text is 100 minus NPV in the BD view. Truth has three levels (effect-dominated 18.8%, mixed 28.7%, bias-dominated 52.5%), and the mixed zone is negative in both views, so the views are not mirror images and no pair of columns sums to 100.

| Strategy | Declare usable (%) | Sens, ED (%) | Spec, ED (%) | PPV, ED (%) | NPV, ED (%) | Sens, BD (%) | Spec, BD (%) | PPV, BD (%) | NPV, BD (%) |
| --- | --- | --- | --- | --- | --- | --- | --- | --- | --- |
| R0 uncalibrated P < .05 (no bias correction) | 67.7 | 82.3 | 35.7 | 22.8 | 89.7 | 31.1 | 66.4 | 50.5 | 46.6 |
| R1 calibrated P < .05 (layer 1 only) | 61.8 | 88.1 | 44.2 | 26.7 | 94.2 | 57.6 | 83.3 | 79.2 | 64.0 |
| R2 layer 1, plus BAF point estimate below 0.5, the bias-dominated threshold | 44.2 | 88.1 | 66.0 | 37.4 | 96.0 | 86.1 | 77.7 | 81.0 | 83.5 |
| R3 layer 1, plus the whole credible interval of BAF below 0.5, the bias-dominated threshold | 35.5 | 84.6 | 75.9 | 44.7 | 95.5 | 93.1 | 67.1 | 75.8 | 89.8 |
| R4 layer 1, plus probability of bias dominance below 15% | 22.1 | 82.1 | 91.8 | 69.7 | 95.7 | 98.7 | 45.1 | 66.5 | 97.0 |
| C1 BAF point estimate below 0.5, the bias-dominated threshold, no layer 1 | 51.3 | 98.3 | 59.6 | 35.9 | 99.3 | 81.7 | 87.8 | 88.1 | 81.3 |
| C2 the whole credible interval of BAF below 0.5, the bias-dominated threshold, no layer 1 | 38.7 | 91.0 | 73.4 | 44.1 | 97.2 | 91.9 | 72.4 | 78.6 | 89.0 |

#### S3.4 Decision flow by stage

Table S5. Decision flow across layers of the rule.

| Stage | N | Bias.dominated..n. | BD.share…. |
| --- | --- | --- | --- |
| all repetitions | 1920000 | 1008000 | 52.5 |
| layer 1 pass (calibrated p < .05) | 1187378 | 427641 | 36.0 |
| layer 1 pass and usable verdict | 424079 | 12804 | 3.0 |
| layer 1 pass and mixed verdict | 272277 | 55739 | 20.5 |
| layer 1 pass and competitive verdict | 132047 | 59781 | 45.3 |
| layer 1 pass and not-usable verdict | 358975 | 299317 | 83.4 |

#### S3.5 Verdict versus truth (3 by 3 confusion)

Table S6. Confusion matrix of assigned verdict zone against simulated truth.

| Truth | bias.dominated | mixed | effect.dominated | Row.total |
| --- | --- | --- | --- | --- |
| bias-dominated | 636531 | 289473 | 81996 | 1008000 |
| effect-dominated | 2486 | 29910 | 327604 | 360000 |
| mixed | 63677 | 155385 | 332938 | 552000 |

Zone accuracy was 58.3% overall, 63.1% among bias-dominated, 28.1% among mixed, and 91.0% among effect-dominated repetitions.

#### S3.6 Subgroup robustness

Table S7. Misuse rate across 7 pre-specified subgroups.

| Subgroup | N | Layer1.pass…. | BD.among.pass…. | Declare.usable…. | BD.among.usable…. | Median.half.width |
| --- | --- | --- | --- | --- | --- | --- |
| K = 12 negative controls | 640000 | 62.4 | 36.3 | 22.4 | 3.46 | 0.144 |
| K = 25 negative controls | 640000 | 61.8 | 35.9 | 22.1 | 3.00 | 0.091 |
| K = 50 negative controls | 640000 | 61.3 | 35.8 | 21.8 | 2.59 | 0.060 |
| bias exchangeability held | 960000 | 60.4 | 35.5 | 19.9 | 1.39 | 0.090 |
| 30% exchangeability violation | 960000 | 63.2 | 36.6 | 24.2 | 4.36 | 0.087 |
| sigma_ps = 0.06 | 960000 | 69.9 | 38.4 | 23.0 | 2.09 | 0.089 |
| sigma_ps = 0.10 | 960000 | 53.8 | 32.9 | 21.2 | 4.02 | 0.088 |

#### S3.7 Discrimination

Table S8. Discrimination (AUC). The area under the curve is read from the bias-dominated view, meaning that withholding a repetition is a positive call and being truly bias-dominated is a positive truth. Figure 7 of the main text reports the same three scores from both viewpoints.

| Predictor | AUC | LR.chi2.note |
| --- | --- | --- |
| calibrated p-value alone | 0.787 | - |
| BAF_hat alone | 0.922 | - |
| BAF_hat + CI half-width | 0.923 | LR chi2 = 3835.1, df = 1, p < .001 |
| BAF_hat + CI half-width + calibrated p | 0.923 | - |

The likelihood-ratio chi-square test for adding credible-interval half-width to BAF̂ was χ² = 3835.1, df = 1, p < .001. The odds ratio per unit increase in CI half-width was 3.88.

#### S3.8 Credible-interval width and coverage

The median half-width was 0.088 (quartiles 0.040, 0.148). Coverage of the Gibbs posterior 95% credible interval was 55.7% overall, 34.4% for narrow intervals, and 76.9% for wide intervals. Mean half-width by K was 0.156 for K = 12 and 0.101 for K = 25. The calibrated P-value at the 0.05 threshold declared 61.8% of repetitions passable versus 67.7% for the uncalibrated P-value; the pass rate among bias-dominated truth was 42.4%, among mixed 80.1%, and among effect-dominated 88.1%.

### S4. Part III supplementary: case-study estimation

#### S4.1 Negative-control empirical null

The jointly estimated empirical null had center μ̂_B = -0.168 and scale σ̂_B = 0.113; the Shapiro-Wilk test of the 12 negative-control residuals gave p = 0.171, and the calibrated log relative risks of the controls ranged from -0.769 to 0.178.

#### S4.2 Full case estimation

Table S9. Full estimation output for the two clinical questions.

| Question | Uncal.RR | Cal.RR | Cal.P | BAFhat | BAF.lo | BAF.hi | p.bias.dom | Verdict |
| --- | --- | --- | --- | --- | --- | --- | --- | --- |
| Beta-blocker >=50% target dose | 0.831 | 0.983 | 0.8940 | 0.875 | 0.385 | 0.992 | 0.999 | not usable as effect evidence |
| GDMT >=2 of 3 classes | 0.682 | 0.807 | 0.1026 | 0.460 | 0.186 | 0.619 | 0.588 | competitive, no verdict |

#### S4.3 Sensitivity analyses (question 1)

Leave-one-out across the 12 negative controls gave BAF̂ ranging from 0.815 to 0.955 (fall-back BAF̂ = 0.941). Replacement of the Super Learner library gave BAF̂ = 0.871 (0.391 to 0.991), 0.871 (0.395 to 0.992), and 0.876 (0.374 to 0.994) under the three alternative libraries. The Gaussian-process propensity-score perturbation gave calibrated relative risk 0.972 to 1.025 and BAF̂ 0.857 to 0.951. All sensitivity analyses preserved the “not usable as effect evidence” verdict for question 1 and the “competitive, no verdict” zone for question 2.

### S5. Additional sensitivity and validation analyses

#### S5.1 Held-out validation of the lookup table

The lookup is built from all 1,920,000 repetitions of the pooled design and then used to score the reporting rules on the same repetitions, so its reported operating characteristics are in-sample. We bounded the resulting optimism by rebuilding the entire lookup, including the bin edges, the median half-width that separates narrow from wide intervals, and the per-bin probabilities, on training data alone, and then applying it unchanged to held-out data. Two schemes were used, each repeated across five folds. In the first, 20% of repetitions were held out within every condition, so that both training and test data span the whole factorial grid; the misuse rate of every rule was then unchanged to two decimal places, which supports the argument that at least 1,000 repetitions per cell makes the in-sample optimism negligible when only repetitions are swapped. In the second, 20% of the 1920 conditions were held out entirely, which additionally tests grid cells that never contributed to the lookup; the share declared usable was essentially unchanged, and the misuse rate of the strictest rule (R4) rose from 2.7% to 2.8%, the largest excursion across the five folds being 4.0% and the largest increase for any rule 0.12 percentage points (Table S10). Generalisation to unseen cells therefore costs little, but it is not free, and the ordering of the rules is preserved.

Table S10. Five-fold held-out validation of the BAF reliability lookup, for the seven reporting rules of Table 4. The lookup, including its bin edges, its median half-width, and its per-bin probabilities, was rebuilt on training data only and applied unchanged to the held-out data; the training columns report its performance on the data used to build it, so the gap between the training and test columns is the in-sample optimism. Misuse is the share of repetitions declared usable that were in truth bias-dominated.

| Held out | Rule | Declared usable, training (%) | Declared usable, test (%) | Misuse, training (%) | Misuse, test (%) | Misuse across folds (%) |
| --- | --- | --- | --- | --- | --- | --- |
| 20% of repetitions held out | R0 | 67.7% | 67.7% | 53.44% | 53.44% | 53.3–53.6 |
| 20% of repetitions held out | R1 | 61.8% | 61.8% | 36.02% | 36.02% | 35.9–36.1 |
| 20% of repetitions held out | R2 | 44.2% | 44.2% | 16.48% | 16.48% | 16.3–16.6 |
| 20% of repetitions held out | R3 | 35.5% | 35.5% | 10.16% | 10.16% | 10.1–10.3 |
| 20% of repetitions held out | R4 | 22.1% | 22.1% | 3.02% | 3.02% | 2.9–3.1 |
| 20% of repetitions held out | C1 | 51.3% | 51.3% | 18.69% | 18.69% | 18.5–18.8 |
| 20% of repetitions held out | C2 | 38.7% | 38.7% | 11.04% | 11.04% | 11.0–11.1 |
| 20% of conditions held out | R0 | 67.7% | 67.7% | 53.44% | 53.47% | 50.5–55.2 |
| 20% of conditions held out | R1 | 61.8% | 61.8% | 36.02% | 36.03% | 32.0–38.4 |
| 20% of conditions held out | R2 | 44.2% | 44.2% | 16.48% | 16.52% | 14.5–18.6 |
| 20% of conditions held out | R3 | 35.5% | 35.5% | 10.16% | 10.24% | 8.1–12.7 |
| 20% of conditions held out | R4 | 20.7% | 20.4% | 2.68% | 2.80% | 0.8–4.0 |
| 20% of conditions held out | C1 | 51.3% | 51.3% | 18.70% | 18.73% | 16.9–21.6 |
| 20% of conditions held out | C2 | 38.7% | 38.7% | 11.05% | 11.14% | 9.1–14.0 |

#### S5.2 Joint propagation of empirical-null and target-estimate uncertainty

The reported credible interval propagates draws of the empirical-null mean while holding the observed target estimate fixed, so it omits the sampling variability of that estimate and undercovers. To quantify what this omission costs, we recomputed the interval with the omitted term added, drawing the calibrated target estimate from a normal distribution centred on its observed value with its own standard error. Both intervals were computed within the same repetitions, so the comparison is paired. Across the settings examined, coverage rose from 78.8% to 97.7%, the median half-width widened from 0.132 to 0.222, and the yield of the strictest interval rule fell from 24.3% to 13.2% of repetitions, with 11.2% of repetitions changing that rule’s decision (Table S11). The undercoverage reported in the main text is therefore attributable specifically to the omitted term. The direction in which that omission propagates to the reporting rules is easy to misstate, and the natural reading is the wrong one. R3 declares an estimate usable only when the whole credible interval lies below 0.5, and a narrower interval satisfies that condition more often, not less often; the reported interval therefore inflates the yield of the rule rather than deflating it. The same argument applies to the misuse rate, because the repetitions that cease to qualify once coverage is restored are those carrying the least evidence against bias dominance: restoring nominal coverage lowered the yield from 24.3% to 13.2% and the misuse rate from 5.1% to 0.6%. The yield and the misuse rate reported for R3 in the main text are accordingly upper bounds on the values that would obtain at nominal coverage, not lower bounds.

Table S11. Interval coverage, precision, and rule performance when the sampling variability of the calibrated target estimate is propagated jointly with that of the empirical null, compared with the reported interval, which holds the target estimate fixed. Both intervals were computed within the same 64,000 repetitions, so the comparison is paired. Rule is the strictest interval rule (R3). The joint interval is wider and attains nominal coverage, at the cost of a lower yield.

| Scope | N | Coverage, reported (%) | Coverage, joint (%) | Median half-width, reported | Median half-width, joint | Rule yield, reported (%) | Rule yield, joint (%) | Rule misuse, reported (%) | Rule misuse, joint (%) |
| --- | --- | --- | --- | --- | --- | --- | --- | --- | --- |
| all retained conditions (K = 12 and 50, exV = 0) | 64,000 | 78.8% | 97.7% | 0.132 | 0.222 | 24.3% | 13.2% | 5.10% | 0.57% |
| K = 12 | 32,000 | 92.7% | 98.5% | 0.185 | 0.247 | 15.7% | 9.2% | 2.69% | 0.24% |
| K = 50 | 32,000 | 64.9% | 96.8% | 0.077 | 0.191 | 33.0% | 17.1% | 6.24% | 0.75% |
| sigma_psi = 0.06 | 32,000 | 85.6% | 98.3% | 0.135 | 0.194 | 23.2% | 15.8% | 1.98% | 0.28% |
| sigma_psi = 0.10 | 32,000 | 72.1% | 97.0% | 0.129 | 0.246 | 25.5% | 10.6% | 7.95% | 1.01% |

#### S5.3 Missing-data handling for the guideline-directed medical therapy exposure

Of the 14,677 patients in the question-2 cohort, 3,494 (23.8%) had an indeterminate guideline-directed medical therapy score at baseline. The primary analysis classified them as not optimized, which is a design decision rather than a data fact. We therefore repeated the question-2 estimation under four conventions: indeterminate as suboptimal (the published analysis), complete case, indeterminate as optimized, and multiple imputation, in which the exposure was drawn from a logistic model of exposure on the ten baseline covariates and combined across ten imputations by Rubin’s rules. The target estimate moved substantially across conventions (uncalibrated relative risk 0.68 to 0.91), but the calibrated P value never fell below .05 and no convention produced a usable determination; the bias fraction rose from 0.460 under the published convention to between 0.463 and 0.676 under the others, and 2 of the 3 alternative conventions moved the determination to not usable as effect evidence rather than the competitive determination returned by the published convention (Table S12). The published convention is thus the most favourable of the four, so the qualitative conclusion is not an artefact of it. The negative-control panel is fitted on the guideline-restricted cohort, so it shares the exposure contrast of the target estimates rather than the contrast that counts all beta-blocker agents. Aligning it in that way changed the empirical null (SD 0.068 under the earlier all-agent panel versus 0.113 here) and moved the question-2 calibrated P value from 0.045 to 0.103, so the layer-1 pass that question 2 would have obtained under the earlier panel was not robust to the realignment; with the aligned panel neither question passes layer 1, while both determinations are unchanged.

Table S12. Question 2 under four conventions for the 3,494 patients (23.8%) with an indeterminate guideline-directed medical therapy score. The bias fraction and its credible interval are computed with the same bootstrap estimator and n_boot used for the published analysis, and the determination is read from the same lookup table, so the only quantity that varies across rows is the target estimate. The first row reproduces the published question-2 result exactly.

| Indeterminate exposure handled as | N | Uncalibrated RR | Uncalibrated RR 95% CI | Calibrated P | BAF | BAF 95% credible interval | P(bias-dominated) | Determination |
| --- | --- | --- | --- | --- | --- | --- | --- | --- |
| indeterminate as suboptimal (published) | 14,677 | 0.682 | 0.597–0.779 | .1026 | 0.460 | 0.186–0.619 | 0.502 | competitive, no verdict |
| complete case (indeterminate dropped) | 11,183 | 0.684 | 0.602–0.777 | .1040 | 0.463 | 0.187–0.623 | 0.502 | competitive, no verdict |
| indeterminate as optimized (opposite extreme) | 14,677 | 0.912 | 0.849–0.979 | .5258 | 0.676 | 0.616–0.933 | 0.976 | not usable as effect evidence |
| multiple imputation (m = 10, Rubin) | 14,677 | 0.745 | 0.653–0.851 | .3377 | 0.598 | 0.242–0.805 | 0.928 | not usable as effect evidence |

#### S5.4 Screening flow for the negative control panel

The four screening levels described in S1.3 are set out in Table S13.

Table S13. Screening flow from the candidate list to the reported negative control panel. Both quantitative thresholds (50 hospitalizations, then 50 events inside the analysis window) were fixed before screening and applied mechanically, and no candidate was removed on the basis of the strength or direction of its estimated association with the exposure.

| Level | Criterion | Entering | Removed | Remaining | Outcomes removed |
| --- | --- | --- | --- | --- | --- |
| Candidate list | Outcomes with no plausible causal pathway from either treatment strategy | 20 | 4 | 16 | Fracture, burn, poisoning, inguinal hernia (no records in the diagnoses table) |
| Code feasibility | Code family represented in the MIMIC-IV diagnoses table | 16 | 0 | 14 | None |
| Cohort event count | At least 50 hospitalizations coded for the outcome in the heart-failure cohort | 14 | 2 | 12 | Conjunctivitis; Non-Melanoma Skin Cancer (44 and 38 qualifying patients) |
| Analysis-window event count | At least 50 events among patients who survived the 7-day grace period | 14 | 2 | 12 | Dental Caries; Varicose Veins (45 and 36 events in the window) |

#### S5.5 Ejection-fraction proxy sensitivity analysis

Eligibility for the case study was anchored on a heart-failure discharge diagnosis rather than on measured left ventricular function, because neither MIMIC-IV nor MIMIC-IV-ECG records the left ventricular ejection fraction, so no ejection-fraction threshold could be applied and the cohort mixes reduced with preserved systolic function (S1.6). To test whether that heterogeneity drives the case-study result, we applied a prespecified proxy.

Probable heart failure with reduced ejection fraction was defined as a systolic heart failure code (ICD-10-CM I50.2x) or an electrocardiographic pattern of QRS duration of at least 120 ms together with left bundle-branch block, a pattern that would satisfy a criterion for cardiac resynchronization therapy and is therefore typical of reduced ejection fraction. Probable heart failure with preserved ejection fraction was defined as a diastolic code (I50.3x) without a systolic code and without that pattern. The two categories are mutually exclusive and together cover only part of the cohort: of 15,053 hospitalizations, 3,367 (22.4%) were classified as probable reduced and 2,535 (16.8%) as probable preserved, leaving 60.8% that met neither definition.

Question 1 was then re-estimated within each proxy subgroup under otherwise identical settings, meaning the same cohort definition, the same ten baseline covariates, the same 7-day grace period, the same estimators, and the same guideline-matched empirical null. The uncalibrated relative risk was 0.813 in probable reduced ejection fraction versus 0.831 in the full cohort; the bias fraction and its credible interval were 0.841 (0.342 to 0.993) against 0.875 (0.385 to 0.992), and the calibrated P value was 0.807 against 0.894. In probable preserved ejection fraction the corresponding values were 0.896, 0.720 (0.629 to 0.973), and 0.774. No subgroup passed layer 1, and all three remained bias-dominated, so the case-study conclusion does not depend on how the ejection fraction is distributed within the cohort. Table S14 gives the full comparison.

Table S14. Question 1 re-estimated within ejection-fraction proxy subgroups under otherwise identical settings (same cohort definition, same 10 baseline covariates, same 7-day grace period, same estimators, same guideline-matched empirical null). Probable reduced ejection fraction is a systolic heart-failure code (ICD-10-CM I50.2x) or QRS of at least 120 ms with left bundle-branch block; probable preserved ejection fraction is a diastolic code (I50.3x) without a systolic code and without that pattern. The two strata are mutually exclusive; 60.8% of hospitalizations met neither definition and are excluded from both.

| Subgroup | N | Deaths, No. | Uncalibrated RR | Calibrated RR | Calibrated P | BAFhat | BAF 95% credible interval | P(bias-dominated) |
| --- | --- | --- | --- | --- | --- | --- | --- | --- |
| Full cohort (all I50.x) | 14,677 | 2,781 | 0.831 | 0.983 | .8938 | 0.875 | 0.385–0.992 | 0.994 |
| Probable HFrEF | 3,278 | 528 | 0.813 | 0.961 | .8071 | 0.841 | 0.342–0.993 | 0.994 |
| Probable HFpEF | 2,504 | 347 | 0.896 | 1.060 | .7741 | 0.720 | 0.629–0.973 | 0.976 |

#### S5.6 Directional coverage of the simulation grid

The BAF definition uses absolute values, which makes the metric indifferent to the direction in which the bias acts, so one BAF value is compatible with bias that inflates the apparent effect, bias that shrinks it toward the null, and bias that reverses its sign. The primary grid sets ψ and μB to the same sign and therefore characterizes only the first of those three roles. To cover the other two, we ran a mirror grid that keeps the ten ψ levels and the eight μB magnitudes of the primary grid but gives μB a positive sign, holding σψ, K, and the exchangeability factor at the same levels, so that the two grids differ in one factor only. Within the mirror grid, the sign of the expected observed effect, ψ + μB, separates the cells into three roles: shrinkage, when μB opposes ψ and is smaller in absolute value; cancellation, when the two are equal in absolute value; and reversal, when μB is larger. The four roles together span 1920 conditions and 1,920,000 repetitions. These are the same repetitions that every performance number in the main manuscript pools; this section disaggregates them by role.

Table S15 reports, for each role, the number of conditions it contains, the mean signed difference between BAF̂ and BAF_true, its root mean squared error, the median half-width of the credible interval, and the coverage of that interval. Table S16 reports, for the same four roles, agreement between the classified zone and the true zone, the share of repetitions that pass layer 1, the yield of the reporting rule R3, the share of the estimates R3 endorses that are in fact bias-dominated, and the share of cells whose true BAF lies above 0.5.

The point accuracy of the estimator carries across roles. The mean signed error of BAF̂ never exceeded 0.069 in absolute value in any role, and the root mean squared error stayed between 0.089 and 0.155. That is the consequence of a definition that depends on the absolute magnitudes of the bias and of the calibrated effect rather than on their signs.

Coverage of the credible interval does not carry across roles. It was 71.1% where the bias inflates the apparent effect, 46.9% where it shrinks the effect toward the null, 18.4% where the two cancel, and 36.1% where the bias reverses the sign of the effect, against a nominal 95% in every role. The difference is not a consequence of where the true value falls on the 0 to 1 scale. Restricting attention to cells whose true BAF lies between 0.35 and 0.45 with K = 50 and σψ = 0.06, coverage was 63.5% under inflation and 24.5% under shrinkage. What moves is the width of the reported interval rather than the error of the estimate: in that matched set the mean of the per-condition median half-widths was 1.02 times the mean root mean squared error under inflation and 0.27 times under shrinkage. The reason is arithmetic. A draw of the bias center that is larger in absolute value makes the calibrated effect smaller whenever the two have the same sign, so the numerator and the denominator of the ratio |μ̂B| / |ψ̃| move in opposite directions and the ratio disperses; when the signs are opposed they move together and the ratio is stable. Bias that opposes the effect therefore returns a narrower interval, and an estimate that looks more precise, than the error of that estimate supports.

The reporting rule R3 reads the upper end of that interval and inherits the shortfall. It endorsed an estimate in 29.8% of repetitions where the bias inflates the effect, 79.9% where it shrinks the effect, 30.3% where the two cancel, and 9.5% where the bias reverses the sign. Zone agreement moves in the opposite direction and is highest in the reversal role, at 84.3%, so the three-zone classification identifies bias dominance even in the role where R3 almost never endorses an estimate; the quantity that governs the safety of the rule is the interval, not the zone. Where the central estimate lies below 0.5, a narrower interval satisfies the condition that the whole interval lie below 0.5 more often, which is one reason the yield of R3 is highest in the shrinkage role and lowest in the reversal role, where the central estimate lies above 0.5 and a narrower interval moves that condition further out of reach. The misuse rate is informative only in the inflation role, where cells of both kinds are present, and there 7.4% of the estimates R3 endorsed were in fact bias-dominated. No cell in the shrinkage or cancellation roles has a true BAF above the 0.5 boundary and every cell in the reversal role does, so in those three roles the misuse rate is fixed by construction rather than estimated. In the reversal role that leaves the 9.5% of repetitions in which R3 endorsed an estimate as false endorsements, every one of them.

Cancellation is a limiting case rather than a typical one. It arises only when the bias and the true effect are equal in absolute value, which sets the true BAF to exactly 0.5 by identity, and it accounts for 36 of the 1,920 conditions. Because the true value then falls on the boundary that separates the zones, coverage of the interval and agreement of the zone coincide exactly at 18.4% in that role, so neither figure should be read as a typical-case rate.

Table S15. Point accuracy of the BAF estimator and coverage of its credible interval when the systematic bias plays each of the four possible roles with respect to the true effect. Inflation is the primary grid, in which the bias has the same sign as the true effect; shrinkage, cancellation, and reversal come from a mirror grid that keeps every other design factor unchanged and gives the bias center a positive sign. The role is assigned by the sign of the expected observed effect, ψ + μB. The mean signed difference and the root mean squared error are on the BAF scale, as is the median half-width of the reported 95% credible interval; coverage is the share of repetitions in which that interval contains the true BAF. The four roles together span 1,920 conditions and 1,920,000 repetitions.

| Role | Conditions | Mean BAFhat minus BAFtrue | RMSE | Median interval half-width | Interval coverage |
| --- | --- | --- | --- | --- | --- |
| Inflation | 960 | −0.069 | 0.155 | 0.130 | 71.1% |
| Shrinkage | 420 | 0.030 | 0.089 | 0.037 | 46.9% |
| Cancellation | 36 | 0.004 | 0.133 | 0.039 | 18.4% |
| Reversal | 504 | −0.047 | 0.142 | 0.046 | 36.1% |

Table S16. Classification and rule performance when the systematic bias plays each of the four possible roles. Zone agreement is the share of repetitions in which the three-zone classification matches the true zone; layer 1 is the calibrated P value below .05; R3 declares an estimate usable only when it passes layer 1 and its whole credible interval lies below 0.5; and the misuse rate is the share of the estimates R3 endorses that are in fact bias-dominated. The final column gives the share of cells in each role whose true BAF lies above 0.5, the boundary of the bias-dominated zone; no cell in the shrinkage or cancellation roles exceeds it and every cell in the reversal role does, so the misuse rate is informative only in the inflation role, where both kinds of cell are present.

| Role | Zone agreement | Layer-1 pass | R3 yield | R3 misuse | True BAF above 0.5 |
| --- | --- | --- | --- | --- | --- |
| Inflation | 53.1% | 61.9% | 29.8% | 7.4% | 52.5% |
| Shrinkage | 42.5% | 86.4% | 79.9% | 0.0% | 0.0% |
| Cancellation | 18.4% | 46.6% | 30.3% | 0.0% | 0.0% |
| Reversal | 84.3% | 42.4% | 9.5% | 100.0% | 100.0% |

### S7. Supplementary figures

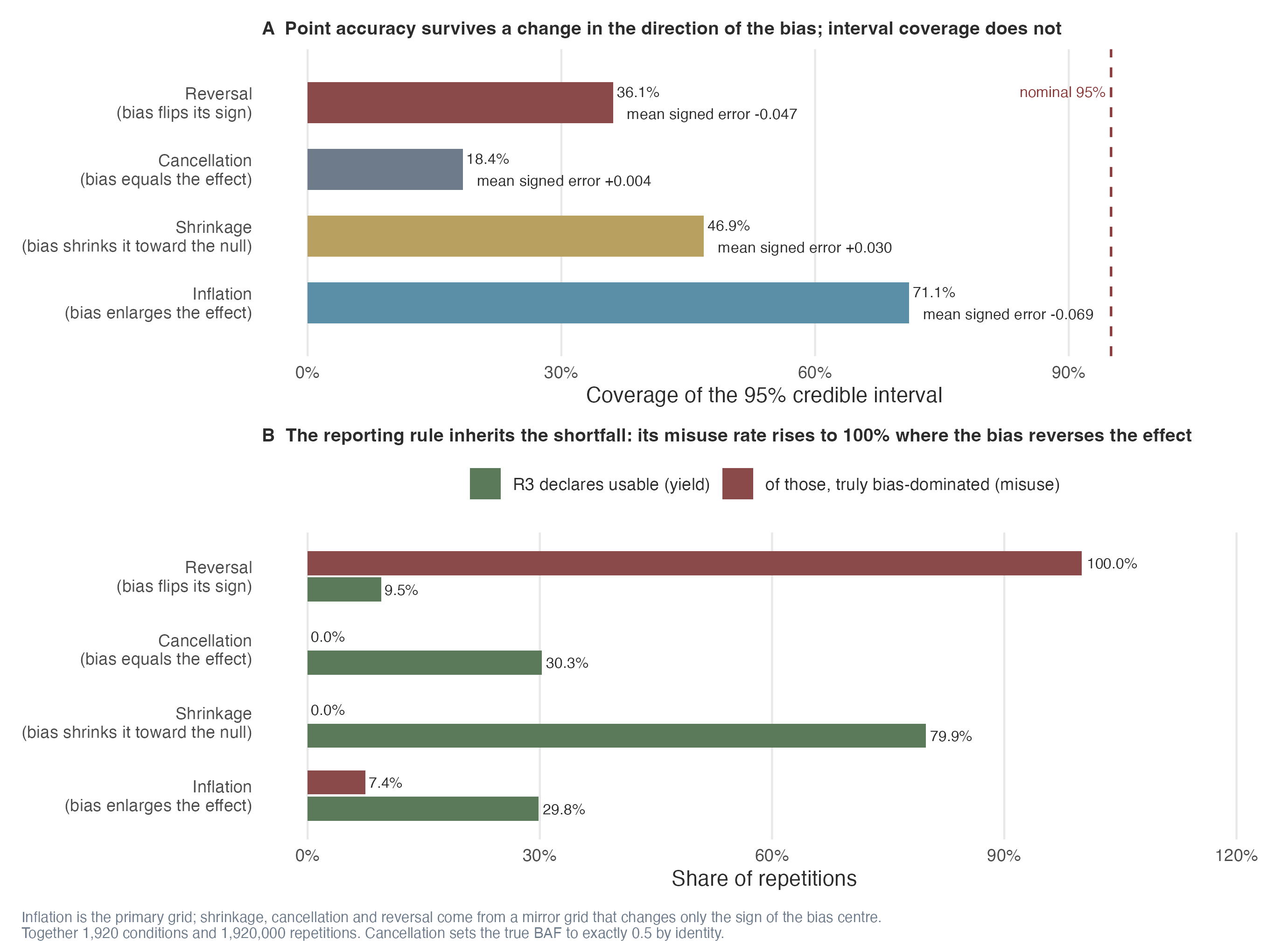

**Figure S1.** Coverage of the 95% credible interval, and performance of the reporting rule R3, when the systematic bias plays each of the four possible roles with respect to the true effect. Panel A gives interval coverage by role, with the nominal 95% level marked; the number beneath each bar is the mean signed error of BAF̂, which stays within 0.069 of zero in every role, so the accuracy of the point estimate carries across roles while the coverage of its interval does not. Panel B gives the share of repetitions R3 declares usable and the share of those that were truly bias-dominated: the rule is at its most permissive where the bias shrinks the effect and at its most dangerous where the bias reverses it, where 100.0% of the estimates it endorses are false endorsements. Both panels use the 1920 conditions and 1,920,000 repetitions of Table S15 and Table S16, and are drawn from the same summary file, so the figure and the two tables cannot drift apart.

### S6. Software and reproducibility

The estimator is implemented in the R package EmpiricalCalibration^1^; the BAF extension is provided by the accompanying R code archived with this manuscript. The analysis pipeline reads MIMIC IV and MIMIC IV ECG under approved data-use agreements; the derived negative-control and estimation tables, the simulation frame, and the single JSON of reported numbers are released with the manuscript repository. The companion interactive explainer consumes the same JSON so that every figure and table in the manuscript and the explainer are guaranteed to agree.

1.	Schuemie MJ, Hripcsak G, Ryan PB, Madigan D, Suchard MA. Empirical confidence interval calibration for population-level effect estimation studies in observational healthcare data. *Proc Natl Acad Sci U S A*. 2018;115(11):2571-2577. doi:[10.1073/pnas.1708282114](https://doi.org/10.1073/pnas.1708282114)
